# JasMAP: A Joint Ancestry and SNP Association Method for a Multi-way Admixed Population

**DOI:** 10.1101/2023.10.26.23297617

**Authors:** Jacquiline Wangui Mugo, Emile Rugamika Chimusa, Nicola Mulder

## Abstract

The large volume of research findings submitted to the GWAS catalog in the last decade is a clear indication of the exponential progress of these studies and association approaches. This success has, however, been dimmed by recurring concerns about disparity and the lack of population diversity. As a result, researchers are now responding, and GWAS extension to diverse populations is under way. Initial GWAS methods were calibrated using European populations with long-range regions of linkage disequilibrium (LD) and haplotypes. This implies that, as GWAS extends to diverse populations, the development of inclusive methods targeted at these populations is imperative. Particularly in multi-way admixed populations, methods that include both genotypes and ancestry associations have been shown to improve power while controlling for the additional LD structure introduced by admixture processes. However, these methods continue to be tailored to only 2-way admixed populations. Though this is a justifiable start, the breeding structures of today suggest that the world population is more likely to increase in the number of multi-admixed individuals, and tools targeted at 2-way admixed individuals will continue to exclude a larger part of diverse populations. In this study, we propose a joint ancestry and SNP association method, JasMAP, that is tailored to multi-way admixed populations. We explore the LMM approach that has become standard in GWAS of structured populations in a Bayesian context, model local ancestry variation as prior knowledge, and update the genotype association to obtain a joint posterior probability of association (PPA). The newly developed method has been assessed using various simulated datasets from our multi-scenario simulation framework, FractalSIM (Mugo et al., 2017), and we output not only the joint statistics but also the genotype-only and the ancestry-only association statistics for the user. JasMAP has also been applied to perform a GWAS analysis of a 5-way admixed South African Coloured (SAC) population with a tuberculosis (TB) phenotype. We obtained 1 significant risk SNP using the ancestry-only association but no SNPs were found to be significant using the standard genotype-only association. 13 risk SNPs, however, were detected as significant with a PPA > 0.5 using the joint association approach. 12 of these SNPs had a marginal significance threshold in genotype-only and ancestry-only association. By functional annotation and gene mapping, the 13 SNPs were found near 8 genes, 5 of which were either found in pathways, have functionality, or were linked to social behaviour associated with an increased risk of TB. Specifically, one of the significant SNPs, *rs*17050321 on chromosome 4, was found close to the *SLC7A11* gene that has previously been linked to TB in a GWAS study of a Chinese population.

## Introduction

The exceptional polygenicity of human traits makes unraveling mechanisms from association studies daunting. In the traditional approach to GWAS, each variant is considered to confer the same risk to the phenotype a priori. This has been explored in numerous methods and has been useful in eliminating potential prior misconceptions, as no preconceived assumptions are made on any region of the genome harboring the disease loci (Shriner et al., 2011). Today, however, researchers are increasingly considering the inclusion of other information in GWAS, and in particular, GWAS studies that include expression quantitative trait loci (eQTLs) are now common (Huang et al., 2015; Li and Kellis, 2016; Thom and Voight, 2020).

The development of disease association studies, their applications for the understanding of disease aetiology, and their evaluation for clinical utility have been underexplored in admixed and diverse populations. In admixed populations, the rich information about the local ancestry effect on the phenotype could be exploited as prior knowledge in a GWAS study and has been shown to improve GWAS power in 2-way admixed populations (Shriner et al., 2011). Harnessing the power of Bayesian approaches to design the next generation of ancestry association models tailored to multi-way admixed populations has the potential to achieve robust methods of genetic association. Therefore, there is a critical need to (1) improve locus ancestry inference (LAI) accuracy; (2) build integrative software for association testing and admixture mapping in multi-way admixed populations; and (3) develop methods for optimizing the predictive power of disease risk in admixed and diverse populations.

The use of LMM approaches for GWAS of structured populations, often in combination with other methods like genomic control, structured association, and principal components (PCs) axes, has over the years been explored and proven to be robust in controlling for a wide range of structures between samples (Kang et al., 2010; Loh et al., 2015; Runcie and Crawford, 2019). In this study, we explore an LMM approach and design a method that jointly models ancestry and SNP association to a phenotype of interest in a multi-way admixed population, called JasMAP. The association analyses within JasMAP will be optimized by combining ancestry and SNP association signals. For robustness and power enhancement, local and global ancestry will be accounted for in a full Bayesian framework. Robust estimation of SNP and ancestry effects will optimize genetic association in multi-way admixed populations.

Our proposed approach within JasMAP extends the BMIX two-step approach to a multi-way admixed population. However, unlike BMIX (Shriner et al., 2011), JasMAP does not require prior knowledge of the parental ancestry with the highest disease prevalence and is targeted at a case-control or quantitative phenotype. It employs the robust LMM approach in a full Bayesian context, and in addition to the joint posterior probability of association summary statistics, it also generates the admixture and genotype association statistics. In the following sections, we discuss the developed method in detail.

## The Proposed Method

The Bayesian inferencing approach is based on the common Bayes theorem, proposed by Thomas Bayes (1701 - 1761). Given two dependent events *A* and *B*, and by defining *𝒫*(*\**) as the probability of event *\** occurring and *𝒫*(*\**_1_|*\**_2_) being the conditional probability of event *\**_1_ occurring given event *\**_2_ has occurred, the Bayes theorem states,

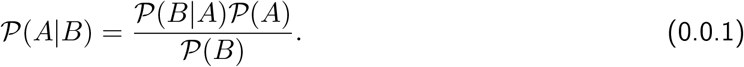

Given the null hypothesis of no association, *H*_0_, and the alternative hypothesis that the SNP is associated with the phenotype, *H*_1_, the posterior probability of the association (PPA) can then be defined as;

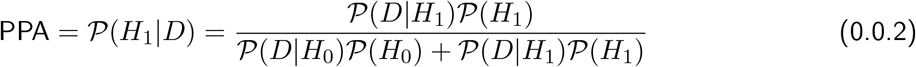

where *𝒫*(*H*_0_) is the prior probability under the null hypothesis, and *𝒫*(*H*_1_) = 1 *− 𝒫*(*H*_0_), is the prior probability under the alternative hypothesis. *𝒫*(*D*|*H*_0_) and *𝒫*(*D*|*H*_1_) are the likelihood functions under the null and alternative hypotheses, respectively.

BMIX considers *𝒫*(*D*|*H*_0_) and *𝒫*(*D*|*H*_1_) as a 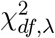, where *df* denotes the degrees of freedom and *λ* is the noncentrality parameter, such that *λ* = 0 for the null hypothesis likelihood function and *λ >* 0 for the alternative hypothesis likelihood function. The p-values can thus be transformed into *χ*^2^ statistics by quantile functions. BMIX further considers a power of 1 *− β* = 0.8, with *β* here denoting the type II error rates. The type I error rate *α* = 0.05. The noncentrality parameter *λ* for the likelihood function under the alternative hypothesis is obtained by the relationship between power and the type I error rate, where for a 1-tailed test,

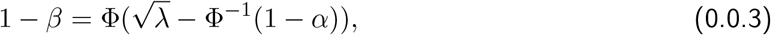

and for a 2 tailed test,

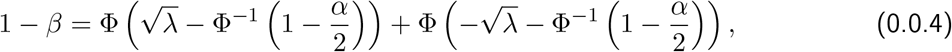

where Φ is the standard Gaussian cumulative distribution function and Φ^*−*1^ is its quantile function.

To account for multiple testing, BMIX employs an autoregressive model to estimate the effective number of tests for the local ancestry and genotypes dataset, which is then used to divide the type I error rate by 0.05 to obtain a partially Bonferroni-corrected significance threshold. In our application, we generated a Python script to obtain the effective number of tests for the ancestry association, which is then provided as input into JasMAP to calculate the partially Bonferroni-corrected threshold. We employ the standard 1.0 *×*10^*−*8^ significance threshold used in GWAS for the genotype association significance threshold if the number of SNPs *≥* 1,000,000; otherwise, we apply Bonferroni correction depending on the number of SNPs in the analysis.

The JasMAP algorithm schematic, **Figure 1** shows a summary of the proposed 2-stage joint association approach. We assume a dichotomous phenotype (case-control) and that the local ancestry inferences are known, as various methods and tools exist to deconvolve local ancestry (Geza et al., 2019). We employ the LMM approach, and in step 1, we perform ancestry association and model the local ancestry inferences and any relevant covariates that affect the association with the phenotype of interest. We also perform genotype association analysis, accounting for any covariates provided, and obtain the corresponding likelihoods. In step 2, we use the summary statistics of the MIA as prior knowledge and the genotype likelihoods from step 1 to obtain our joint association summary statistics. We define the MIA as the ancestry with the lowest p-value from step 1.

**Figure 1.**
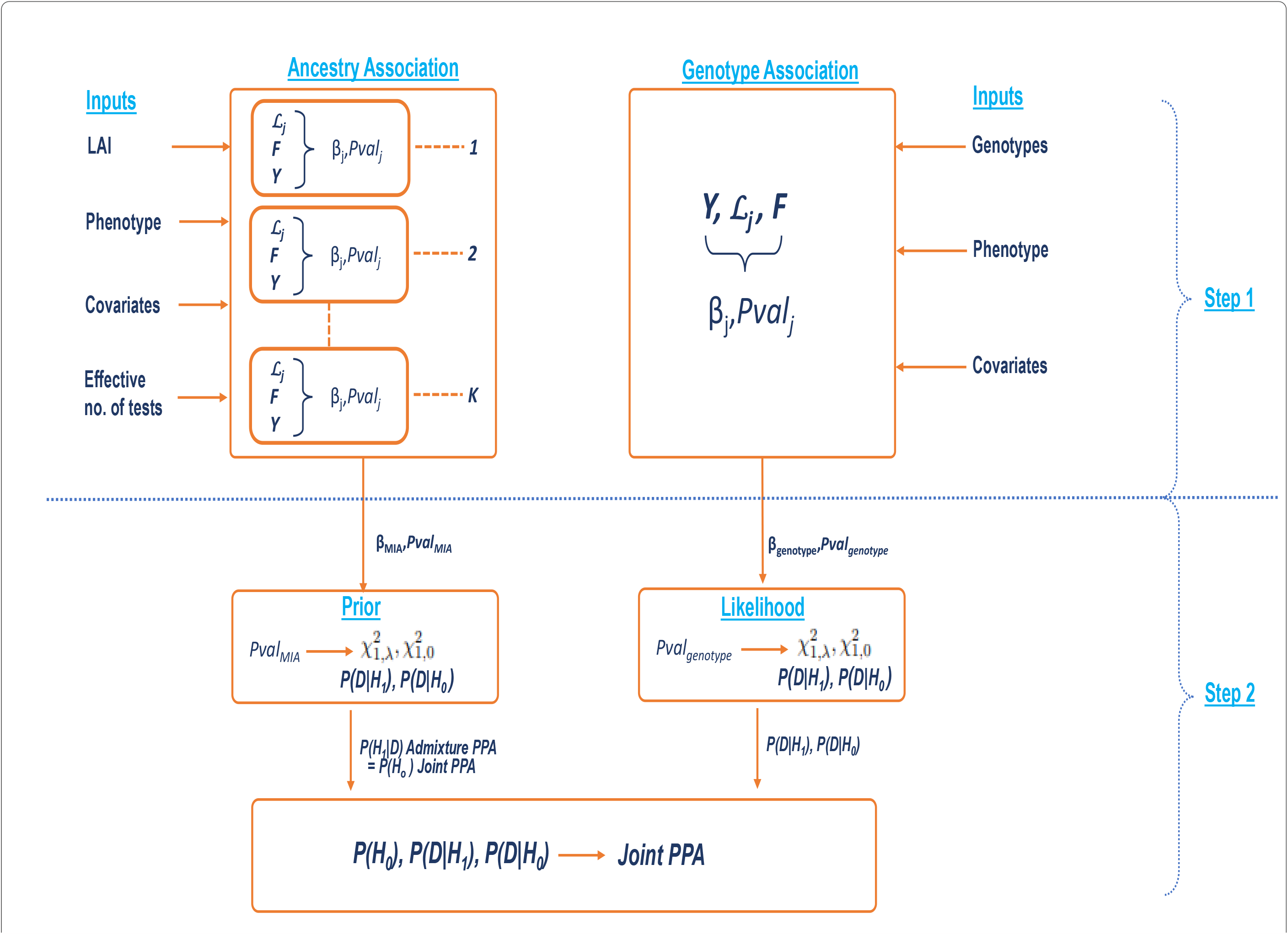
The figure illustrates the JasMAP algorithm’s schematic pipeline. Ancestry association on the left is performed using the local ancestry inferences (LAI), any covariates included, the effective number of tests, and the phenotype information. Genotype association on the right requires as input genotypes, phenotypes, and any available covariates. Both approaches implement LMM to perform the association. The P-value of the most informative ancestry (MIA) is then used as a prior to inform the genotype association in the joint association to obtain the joint posterior probability of the association (PPA).

We describe below each step of the approach and how we obtain the prior, likelihood, and posterior probabilities of association, but first define the notations used and discuss the LMM approach employed in JasMAP.

### Method Notation Definition

- *N* : is the number of admixed individuals considered in the analysis denoted by *i* ∈ {1, 2, 3, …, *N* }.
- *M* : is the number of biallelic SNPs being analyzed, denoted by *j* ∈ {1, 2, 3, …, *M* }.
- *K* : is the number of ancestral populations involved in the admixture process denoted by *k* ∈ {1, 2, 3, …, *K* }.
- *C* : is the number of covariates being considered.
- *Y* : is the *N ×* 1 vector of phenotype, where *Y* ∈ {0, 1} for a dichotomous phenotype, with 0,1 denoting a control and case individual, respectively.
- *L* : is *N* × *M* matrix of standardized data where each entry 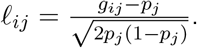.

In genotype association, *L* is the matrix of standardized genotype data, where *g*_*ij*_ *∈* {0, 1, 2} denotes the number of reference alleles at a given SNP *j* for individual *i*, while *p*_*j*_ is the minor allele frequency of SNP *j*.

In ancestry association, *L* is the matrix of standardized local ancestry inferences (LAI) from ancestry *k*, where *g*_*ij*_ *∈* {0, 1, 2} denotes the number of copies of alleles from a specific ancestry at a given SNP *j* for individual *i*, while *p*_*j*_ is the average local ancestry from ancestry *k* at SNP *j* calculated as 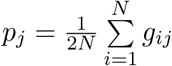.

- *F* : is the *N ×* (*C* + 1) matrix of the covariates, which may include PCs, age, and gender, among others. *F*_0_ : is the intercept term.
- *ω* : is the (*C* + 1) *×* 1 vector of fixed effects of the covariates, where *ω*_0_ is the intercept effect.
- *θ* : is the *M ×* 1 vector of fixed effect sizes, corresponding to the SNP effect in genotype association or the ancestry effect in ancestry association.
- *Z* : is the *N × M* design matrix of the random effects.
- *ψ* : is the *M ×* 1 vector of random effect sizes.
- *ξ* : is the *N ×* 1 vector of residual errors.
- *G* and *E* : are the variances of the random and residual effects, respectively.
- 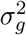 and 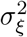: are the variance components corresponding to the random and residual effects, respectively.
- *𝒱* : is the phenotypic variance.
- *𝒦* : is the relatedness matrix or kinship matrix.
- *ℐ* : is the identity matrix.

### Linear Mixed Model Implemented in JasMAP

Let *X* be the combined *N ×* (*M* + *C* + 1) matrix of elements in matrix *L* and *S*, and *β* the combined (*M* + *C* + 1) *×* 1 vector of fixed effects vector *ω* and *θ*. The LMM implemented in JasMAP is thus defined as:

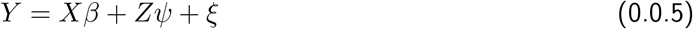

The random and residual effects are assumed to be independent and normally distributed, each with a mean of 0 and variances *G* and *E*, respectively.

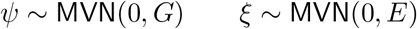

therefore,

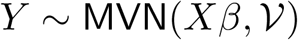

where the phenotypic variance is defined as,

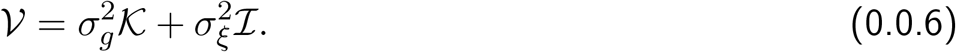

The log likelihood of the phenotype data given the mean and the variance is defined as:

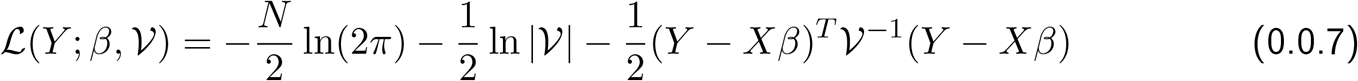

To compute **Equation** 0.0.7, we use an approach first introduced by Kang et al. (2008) in their tool EMMA and more efficiently used by Lippert et al. (2011) in their tool FaST-LMM. Lippert et al. (2011) proposed computation of **Equation** 0.0.7 in linear time. In their algorithm, Lippert et al. (2011) use the parameter

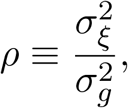

introduced by Kang et al. (2008), referred to as a pseudo-heritability, and thus the phenotypic variance in **Equation** 0.0.6 now becomes,

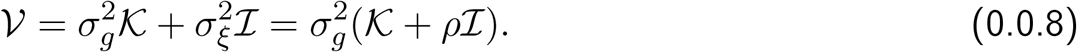

The kinship matrix *𝒦* being a square and symmetric matrix, can be factored into *𝒦* = **Ψ***D***Ψ**^*T*^ by spectral decomposition, where for a full rank matrix *𝒦*, matrix **Ψ** is an *N × N* matrix of eigenvectors of matrix *𝒦*, matrix **Ψ**^*T*^ being the corresponding transpose and *D* a diagonal matrix of the eigenvalues of matrix *𝒦*. By the properties **ΨΨ**^*T*^ = *ℐ*, |**Ψ**| = |**Ψ**^*T*^ | = 1 and **Ψ**^*−*1^ = **Ψ**^*T*^ of the eigenvector matrix **Ψ**, and given that for any two matrices *A* and *B*, |*AB*| = |*A*||*B*|, then **Equation** 0.0.7 can now be written as,

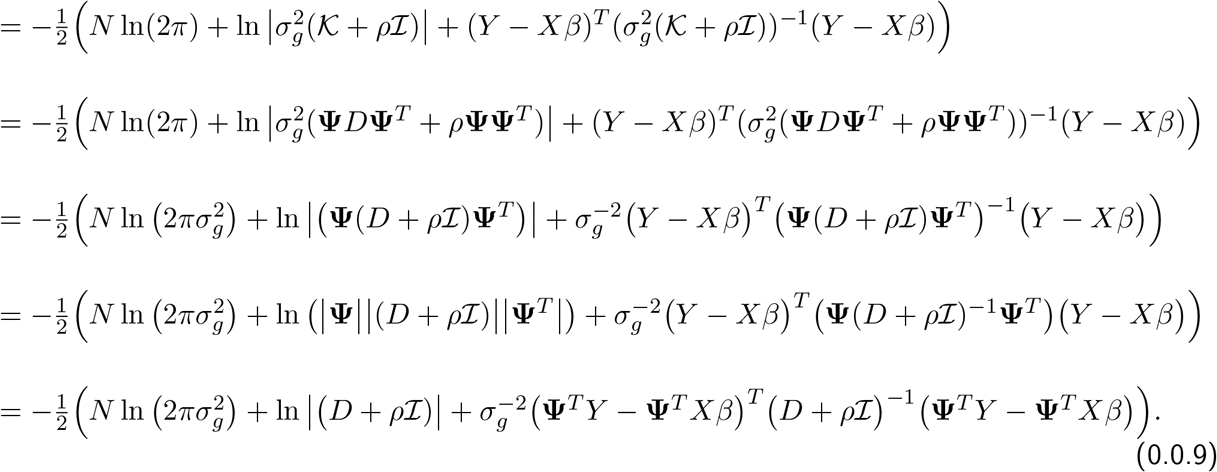

**Equation** 0.0.9 can now be written as,

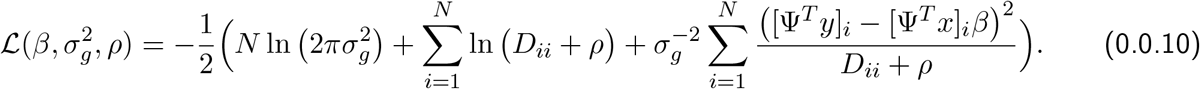

**Equation** 0.0.10 follows from **Equation** 0.0.9 as the covariance matrix, *D* + *ρ ℐ* is diagonal, and thus **Equation** 0.0.9 can be summed over *N*, where *D*_*ii*_ are the entries on the diagonal axis. The computation of the log-likelihood proceeds by first transforming the genotype and phenotype matrices *X* and *Y* to **Ψ**^*T*^ *X* and **Ψ**^*T*^ *Y*, respectively, once, and having estimated the values of *ρ, β* and 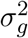, the computation occurs in linear time depending on the sample size *N*.

We obtain the matrix **Ψ** by spectral decomposition of the kinship matrix *𝒦*. This implies that the only unknown parameters in the log-likelihood are *β, ρ* and 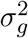. By using maximum likelihood estimation (MLE) to estimate the parameters *β* and 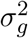, we obtain:

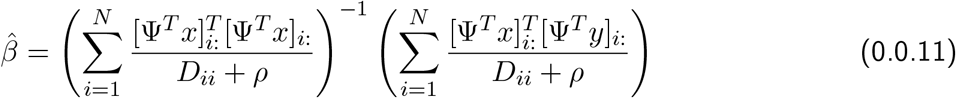

and

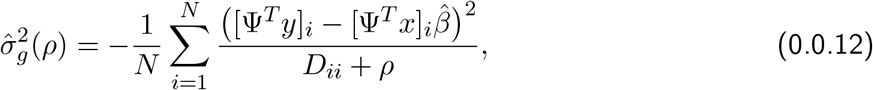

where *i* : is the index of the *i*^th^ row of the matrix. Plugging **Equation** 0.0.11 into 0.0.12, then substituting these values into **Equation** 0.0.10 we get,

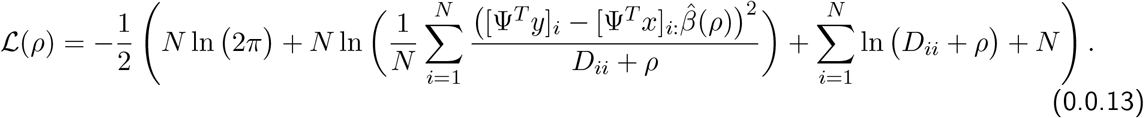

The restricted maximum likelihood (ReML) as suggested by Kang et al. (2008) and also implemented by Lippert et al. (2011) is calculated as,

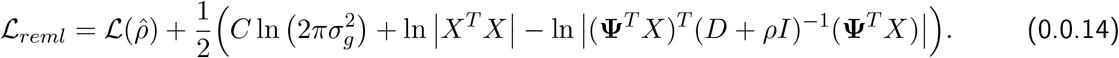

This implies that the task lies in estimating the parameters *ρ*. Similar to Lippert et al. (2011), we optimize **Equation** 0.0.13 to obtain the value of *ρ* that maximizes the log-likelihood function by implementing Brent’s optimization method and obtain the value of *ρ* that maximizes *ℒ*(*ρ*) at each subinterval and store the maximum as 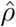. To reduce the computation cost, 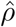 is estimated only once over a null model, 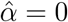, that is, excluding genotype or ancestry effects from the model. This estimate is then used in all the computations to obtain the likelihood.

### The Prior Knowledge: Ancestry Association

In this step, as illustrated in the schematic **Figure 1**, the input includes the local ancestry inference information, any relevant covariates, the phenotype, and the admixture mapping effective number of tests for the admixed population. The MALD explored in the ancestry association extends over a wide region for recently admixed populations (*<* 20 generations), and thus the effective number of tests in the admixture mapping is less than the number of SNPs. A separate script is provided for the user to obtain this input. The script obtains the total local ancestry breakpoints per chromosome per individual, which are averaged across all the individuals in the analysis.

JasMAP accepts local ancestry information as a *M ×* 2*N* matrix. **Figure 2** shows a sample input file for local ancestry inference information for 3 individuals at 4 SNPs in a 3-way admixed population. The rows correspond to the number of SNPs, and the columns correspond to the number of haplotypes, such that two consecutive columns represent local ancestry inferences for a given individual *i* in each of the haplotypes. Each entry on the matrix thus indicates the inferred ancestry at SNP *j* for individual *i* at haplotype 1 or haplotype 2.

**Figure 2.**
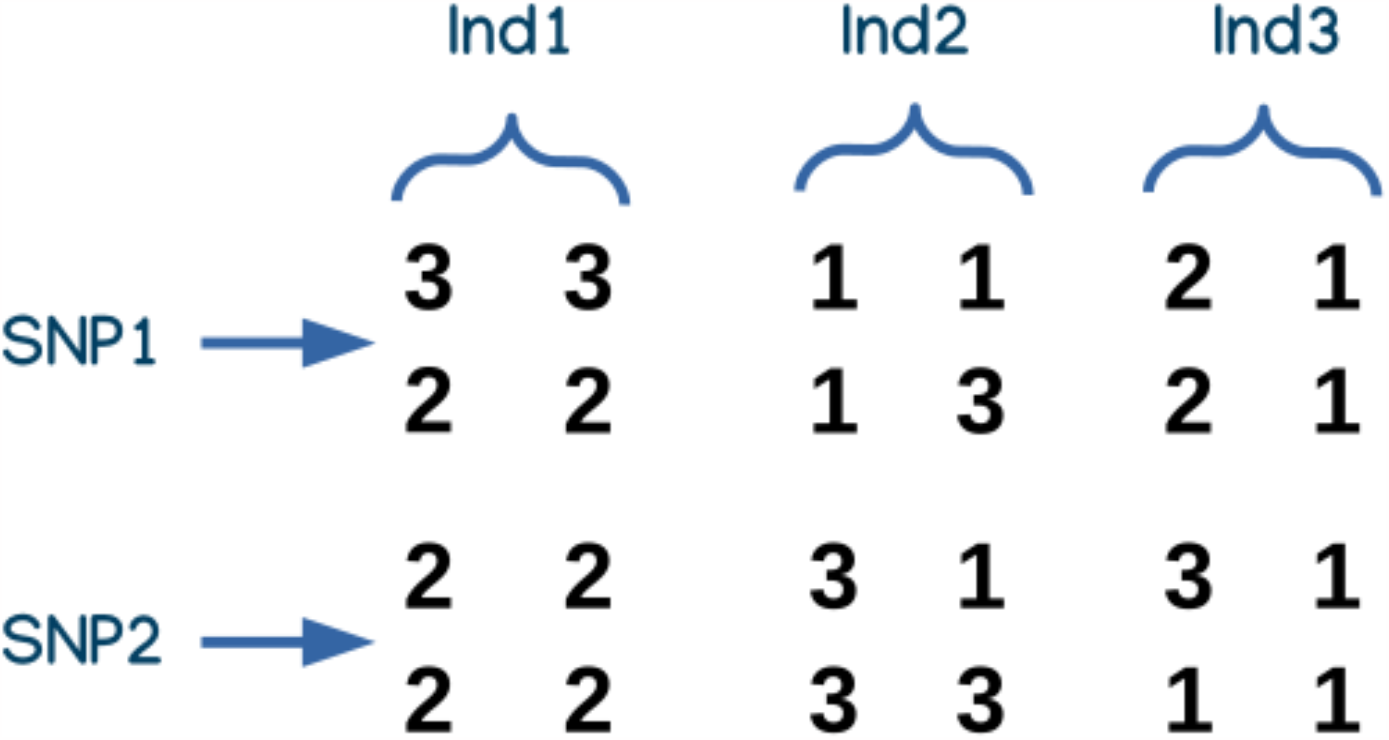
A sample dataset for local ancestry inference file input for JasMAP, with the rows corresponding to the number of SNPs and the columns the number of haplotypes of the individuals in the study. Each entry represents the ancestry inference number *k* ∈ {1, 2, 3} representing the 3 ancestral populations for a specific SNP at a given haplotype. Ind1, Ind2, and Ind3 denote the three individuals.

First, JasMAP obtains the LAI information and splits it into *K*, *L* matrices depending on the number of ancestries, as illustrated in **Figure 2**. For each ancestry, JasMAP then implements the LMM approach by considering only the SNP where the average local ancestry *p*_*j*_ *>* 0.01 and obtaining the corresponding summary statistics.

For each SNP, we obtain the p-values corresponding to the association of each of the ancestries with the phenotype, which is output for the user as the admixture-only association. We then consider the ancestry with the most significant p-value for that SNP, which we call the most informative ancestry (MIA), and convert its corresponding p-value to *χ*^2^ statistics. We then convert the *χ*^2^ statistics to the density functions 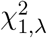 and 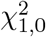, corresponding to *𝒫* (*D*|*H*_1_) and *𝒫* (*D*|*H*_0_) likelihood functions, respectively, where *λ* is calculated. We let the prior probability estimate under the null be *𝒫* (*H*_0_) = 1*/ ν*, where *ν* is the effective number of tests for admixture provided as input by the user, and implementing **Equation** 0.0.2, proceed to obtain the admixture PPA.

### The Likelihood: Genotype Association

JasMAP implements the genotype association per SNP. The input for this step includes the *L* genotypes matrix, the *F* matrix of covariates if provided, and the*Y* phenotype matrix. Similar to ancestry association, JasMAP implements the LMM approach and obtains the p-value corresponding to 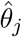 for each SNP *j*. This p-value is then converted to the density functions for genotype association 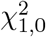 and 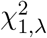 corresponding to the *𝒫*(*D*|*H*_0_) and *𝒫*(*D*|*H*_1_) density functions for the genotype association.

### Joint Posterior Probability of Association

Similar to the SNP association step, we obtain the joint PPA per SNP based on **Equation** 0.0.2. The prior probability *𝒫* (*H*_0_) is the admixture posterior probability of the association while the density functions *𝒫* (*D*|*H*_0_) and *𝒫* (*D*|*H*_1_) are the genotype density functions. We set the standard significance threshold for PPA at 0.5.

## Assessment of JasMAP using Simulated Populations

### Method

We assessed JasMAP using 3-way and 5-way admixed simulated datasets generated under a single-point admixture scenario, where the admixture process occurs at a single point in history, such that the current generation is the offspring of the admixed population that has interbred over the years. Considering a random mating model where interbreeding has occurred for 10 generations, the admixture simulation first mimicked the isolated growth of each population, where a disease model (causal or null) was simulated in the isolated homogeneous simulation for each of the parental populations. At generation 0, the isolated populations were then allowed to interbreed. **Table 1** lists the reference parental populations used in the 2 scenarios, their corresponding initial sample sizes, and the proportion of ancestry contribution of each of the populations.

**Table 1:** The table provides information on the parental reference populations for the 3-way and 5-way admixture simulations, their abbreviations, initial sample sizes, and the percentage of ancestry each population contributed in each scenario.

| Simulation | Parental Population | Sample Size | Ancestry Proportion |
| --- | --- | --- | --- |
| 3-way | Utah residents with Northern and Western European ancestry (CEU) | 94 | 20% |
|  | Han Chinese from Beijing, China (CHB) | 103 | 10% |
|  | Yoruba (YRI) | 108 | 70% |
| 5-way | Europeans (EUR) | 305 | 15% |
|  | South Asians (SAS) | 386 | 35% |
|  | East Asians (EAS) | 441 | 10% |
|  | Other African ancestries (MAFR) | 256 | 10% |
|  | West Africans (WAFR) | 405 | 30% |

In the 3-way simulation, we included 466,142 biallelic SNPs that were present in the 3 parental populations. We simulated four risk SNPs, where we selected one SNP each on chromosomes 2, 6, 11, and 15, and generated 2500 cases and 2500 controls. In the 5-way admixture scenario, we incorporated 623,330 biallelic SNPs that were present in all 5 parental populations and simulated 8 risk SNPs on chromosomes 2, 6, 11, 15, and 20. On chromosomes 2, 11, and 20, we selected two SNPs in each chromosome that were in high LD and one SNP each on chromosomes 6 and 15. In the 5-way scenario, however, we simulated two sets of datasets of different sample sizes: a dataset of 500 cases and 500 controls and another of 2500 cases and 2500 controls.

In the admixture simulation, we simulated different risk scenarios for the different chromosomes by varying the presence and strength of genotype risk on the risk variant simulated and the ancestry risk on the genomic region containing the variant. We simulated ancestry risk by simulating ancestry deviation between cases and controls in the region that contained the risk variants. In the 3-way simulation on chromosomes 2 and 11, we simulated strong genotype and ancestry risks; on chromosome 6, we simulated very strong ancestry risk and weak genotype risk; and on chromosome 15, we simulated weak genotype and ancestry risks. All the other chromosomes were simulated under a null model in this scenario. In the 5-way simulation, we simulated similar levels of risk in the 500 cases and 500 controls and 2,500 cases and 2,500 controls sample sizes. On chromosome 2, we simulated strong genotype and ancestry risks; on chromosomes 6 and 20, we simulated a strong genotype and no ancestry risk; on chromosomes 11 and 15, we simulated weak genotype and ancestry risks; and a null model on all the other chromosomes.

The risk SNPs simulated in the 3-way and 5-way scenarios and their respective homozygosity and heterozygisity relative risks specified for the cases are listed in **Table 2**. Depending on the MAF of the risk SNPs, the specified risks introduced risk signal strength as indicated on **Table 3**.

**Table 2:**
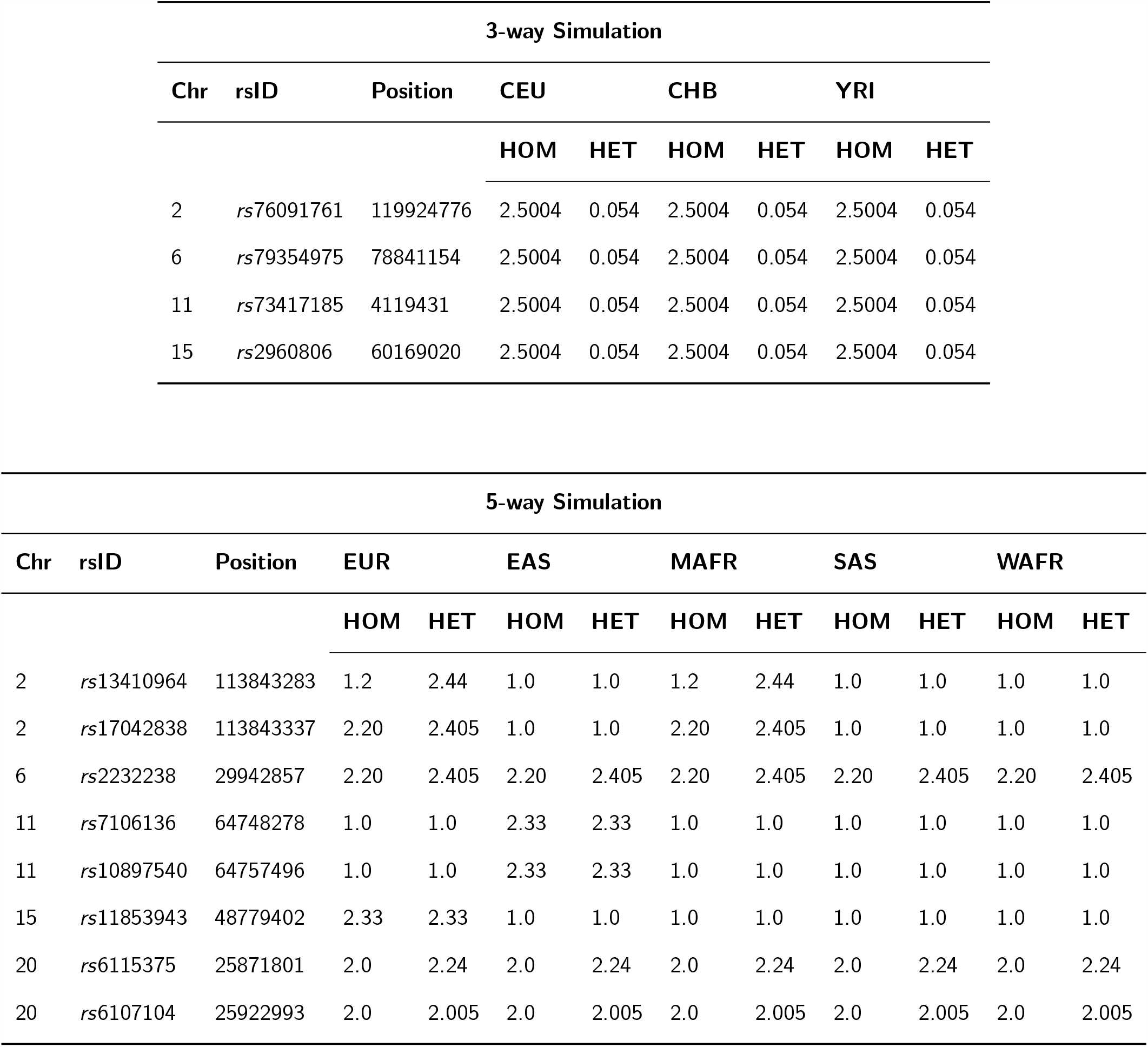
A list of simulated risk SNPs and the corresponding homozygosity (HOM) and heterozygosity (HET) relative risks specified during the isolated simulation of Europeans (EUR), East Asians (EAS), West Africans (WAFR), South Asians (SAS), and other African populations (MAFR) before the admixture process.

**Table 3:** The table lists the disease risk scenarios simulated in the 3-way and 5-way admixture simulations and the chromosomes containing the risk SNP. ✓ indicates a strong risk was simulated, (✓) indicates a weak risk was simulated, while ✗ indicates no risk was simulated.

| Simulation | Chromosome | Genotype Risk | Ancestry Risk |
| --- | --- | --- | --- |
| 3-way | 2 | ✓ | ✓ |
|  | 6 | ✓ | ✓ |
|  | 11 | (✓) | ✓ |
|  | 15 | (✓) | ✓ |
|  | Others | ✗ | ✗ |
| 5-way | 2 | ✓ | (✓) |
|  | 6 & 20 | ✓ | ✗ |
|  | 11 | (✓) | (✓) |
|  | 15 | (✓) | ✓ |
|  | Others | ✗ | ✗ |

We then implemented RFMIX (Maples et al., 2013) tools on these simulated admixed data to obtain the corresponding local ancestry inference matrix input for JasMAP for each set of datasets. Though a number of tools have been developed to deconvolve ancestry in a multi-way admixed population (Geza et al., 2019) and research is still underway to improve these tools, as shown by Geza et al. (2020) in the assessment of the different tools under different scenarios, no tool performs best under all the scenarios. RFMIX was, however, shown to be among the top-performing tools in deconvolving the ancestry of multi-way admixed populations of recently admixed populations and, in addition, has output similar to the input format required by JasMAP. It is important to note, however, that output from other tools can easily be converted to the JasMAP input format.

To ensure a fair assessment of JasMAP, we also obtained the true local ancestry estimates output from FractalSIM during simulation to compare with the RFMIX output. This implies we performed a total of six assessments, two for each dataset: the 3-way and the two sets of 5-way admixed populations. We then implemented a custom Python script and obtained an estimate of the effective number of tests for each of the three simulated datasets, for both the true local ancestry and the RFMIX inferences. **Table 4** shows the effective number of tests obtained for each of the datasets and the corresponding Bonferroni-corrected significance threshold set for each dataset.

**Table 4:** The effective number of tests obtained for the ancestry association for each of the simulated dataset and the corresponding significance threshold set based on the number (AM threshold).

| Simulation | Sample Size | Effective Number of Tests |  | AM threshold |  |
| --- | --- | --- | --- | --- | --- |
|  |  | True | RFMIX | True | RFMIX |
| 3-way | 2500 cases, 2500 controls | 510 | 75 | $9.8039 \times 10^{-05}$ | $6.6667 \times 10^{-04}$ |
| 5-way | 500 cases, 500 controls | 356 | 225 | $1.4045 \times 10^{-04}$ | $2.2222 \times 10^{-04}$ |
| | 2500 cases, 2500 controls | 388 | 211 | $1.2887 \times 10^{-04}$ | $2.3697 \times 10^{-04}$ |

We used 10 PCs as covariates. The LAI estimates, the covariates, the effective number of tests, and the genotypes and phenotypes were then given as inputs to JasMAP. For each assessment, JasMAP outputs the summary statistics corresponding to the MIA per SNP, the genotype association, and the corresponding joint PPA. In addition, JasMAP outputs the ancestry association summary statistics for each of the ancestors. We used this output to plot the corresponding Manhattan plots.

## Results

**Table 5** lists the summary statistics for the simulated risk SNPs for the 3-way admixed simulation while **Figure 3** shows the Manhattan plots corresponding to the 3-way admixed population for the MIA per SNP, the genotype-only association, and the joint PPA for both the true local ancestry and the RFMIX inferences. The Manhattan plots of the ancestry-only association for each of the ancestries used in the 3-way admixture simulation are shown in **Figure 4**, using both the RFMIX inferences and true local ancestry information. **Table 6** lists the summary statistics for the simulated risk SNPs for the smaller sample size 5-way admixed population analysis. **Figure 5** shows the Manhattan plots corresponding to the p-values of the MIA per SNP, the genotype-only association, and the joint PPA for both the true local ancestry and the RFMIX inferences for this dataset. **Figures** 6 and 7 show the Manhattan plots for each ancestry used in the 5-way simulation for the ancestry-only association for both the true local ancestry and RFMIX estimates. **Table 7** lists the summary statistics for the simulated risk SNPs for the larger sample size. The Manhattan plots corresponding to the p-values of the MIA per SNP, the genotype-only association, and the joint PPA for both the true local ancestry and the RFMIX inferences for the larger dataset are shown in **Figure 8**. **Figure 9** and 10 show the corresponding Manhattan plots for each ancestry for the ancestry-only association for both the true local ancestry and RFMIX estimates.

**Table 5:** The list of risk SNPs simulated under the 3-way admixture scenario, with the correponding MIA selected, the p-value under the ancestry association (AM Pval), the genotype association p-value (Genotype Pval), and the PPA for the joint association (Joint PPA) using the RFMIX local ancestry inference (RFMIX) and the true local ancestry (TRUE).

| rsID | CHR | Position | RFMIX |  |  |  | TRUE |  |  |  |
| --- | --- | --- | --- | --- | --- | --- | --- | --- | --- | --- |
|  |  |  | MIA | AM Pval | Genotype Pval | Joint PPA | MIA | AM Pval | Genotype Pval | Joint PPA |
| rs76091761 | 2 | 119924776 | CEU | 0.00208 | $4.69869 \times 10^{-15}$ | 0.9999 | CHB | $1.63152 \times 10^{-25}$ | $4.69869 \times 10^{-15}$ | 0.9999 |
| rs79354975 | 6 | 78841154 | CHB | 0.00133 | $1.82564 \times 10^{-10}$ | 0.9999 | CEU | $1.73069 \times 10^{-100}$ | $1.82564 \times 10^{-10}$ | 0.9999 |
| rs734171851 | 11 | 4119431 | CHB | $6.17746 \times 10^{-06}$ | $7.54124 \times 10^{-05}$ | 0.9999 | 1 | $2.22346 \times 10^{-101}$ | $7.54124 \times 10^{-05}$ | 0.9999 |
| rs2960806 | 15 | 60169020 | 3 | 0.00382 | $1.68258 \times 10^{-06}$ | 0.9997 | YRI | $7.98378 \times 10^{-10}$ | $1.68258 \times 10^{-06}$ | 0.9999 |

**Table 6:** The list of risk SNPs simulated under the 500 cases 500 controls 5-way admixture scenario, with the correponding MIA selected, the p-value under the ancestry association (AM Pval), the genotype association p-value (Genotype Pval), and the PPA for the joint association (Joint PPA) using the RFMIX local ancestry inference (RFMIX) and the true local ancestry (TRUE).

| rsID | CHR | Position | RFMIX |  |  |  | TRUE |  |  |  |
| --- | --- | --- | --- | --- | --- | --- | --- | --- | --- | --- |
|  |  |  | MIA | AM Pval | Genotype Pval | Joint PPA | MIA | AM Pval | Genotype Pval | Joint PPA |
| rs13410964 | 2 | 113843283 | SAS | 0.35931 | $2.0230 \times 10^{-05}$ | $1.3677 \times 10^{-03}$ | WAFR | 0.00179 | $2.02304 \times 10^{-05}$ | 0.9865 |
| rs17042838 | 2 | 113843337 | SAS | 0.35931 | $1.01154 \times 10^{-10}$ | 0.9991 | WAFR | 0.00179 | $1.01154 \times 10^{-10}$ | 0.9999 |
| rs2232238 | 6 | 29942857 | EUR | 0.39185 | $3.86936 \times 10^{-21}$ | 0.9999 | EAS | 0.5108 | $3.86936 \times 10^{-21}$ | 0.9999 |
| rs7106136 | 11 | 64748278 | MAFR | 0.6543 | 0.00071 | $6.8095 \times 10^{-07}$ | MAFR | 0.00174 | 0.00071 | 0.2482 |
| rs10897540 | 11 | 64757496 | MAFR | 0.6543 | 0.00026 | $3.606 \times 10^{-06}$ | MAFR | 0.00174 | 0.00026 | 0.6361 |
| rs11853943 | 15 | 48779402 | MAFR | 0.00997 | 0.00016 | 0.1358 | SAS | $3.50647 \times 10^{-05}$ | 0.00016 | 0.9973 |
| rs6115375 | 20 | 25871801 | MAFR | 0.20729 | $3.32176 \times 10^{-24}$ | 0.9999 | WAFR | 0.55617 | $3.32176 \times 10^{-24}$ | 0.9999 |
| rs6107104 | 20 | 25922993 | MAFR | 0.20729 | $1.29474 \times 10^{-13}$ | 0.9999 | WAFR | 0.55617 | $1.29474 \times 10^{-13}$ | 0.9999 |

**Table 7:**
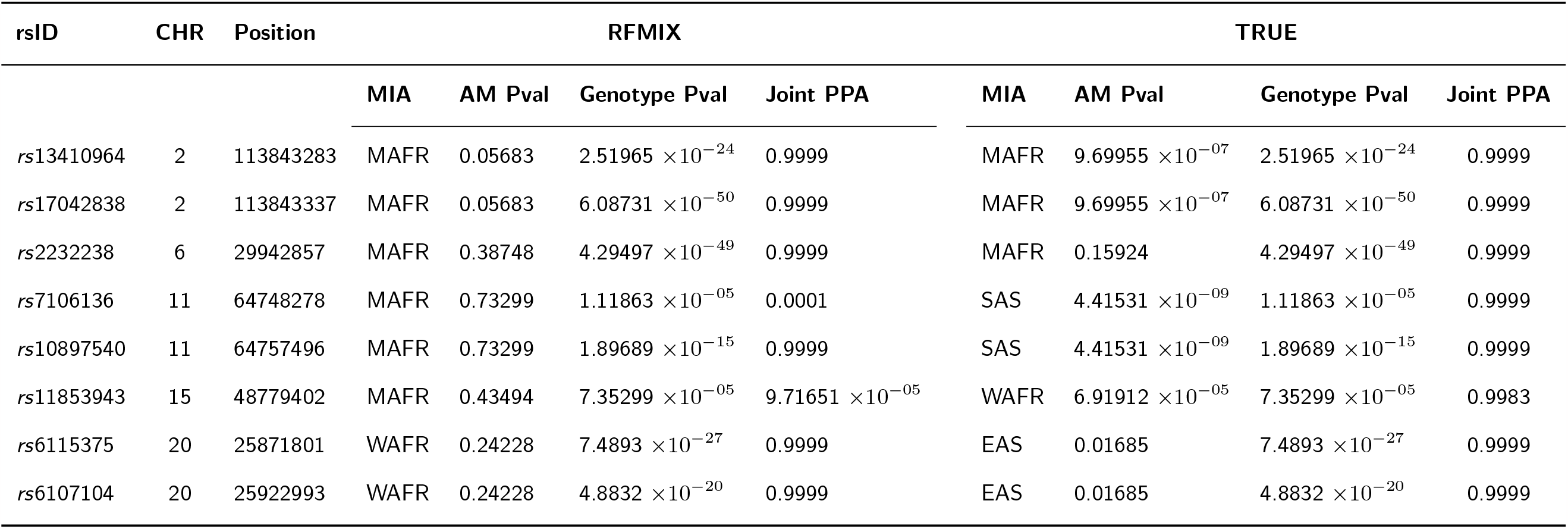
The list of risk SNPs simulated under the 2500 cases 2500 controls 5-way admixture scenario, with the correponding MIA selected, the p-value under the ancestry association (AM Pval), the genotype association p-value (Genotype Pval), and the PPA for the joint association (Joint PPA) using the RFMIX local ancestry inference (RFMIX) and the true local ancestry (TRUE).

**Figure 3.**
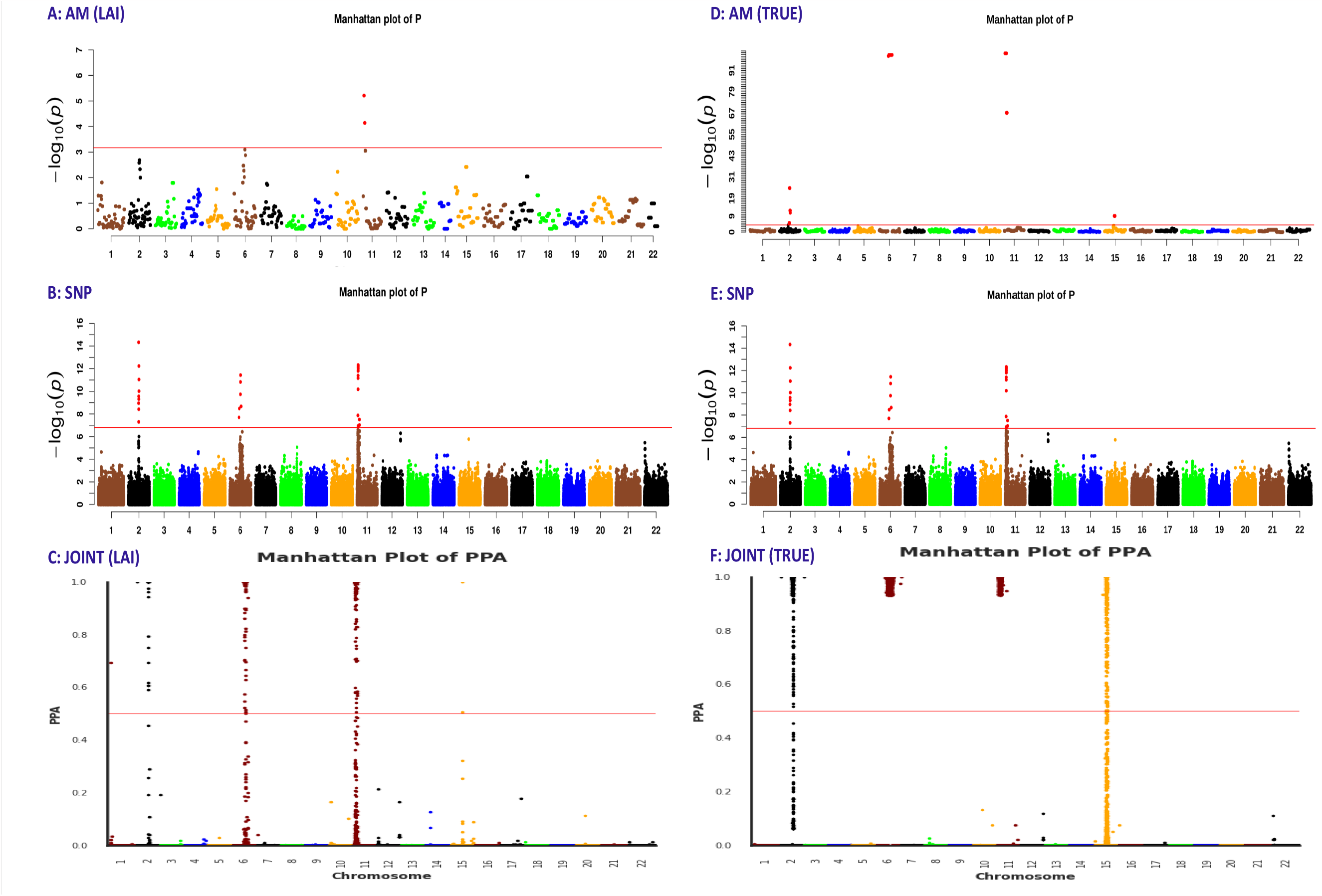
Manhattan plots A, B, and C correspond to the association analysis using the RFMIX local ancestry inferences (LAI), while D, E, and F correspond to the association analysis using the TRUE local ancestry. A and D correspond to the p-values of the MIA per SNP, B and E to the genotype association, and C and F to the joint PPA for the 3-way admixed population analysis. The significance threshold line and the significant SNPs are shown in red.

**Figure 4.**
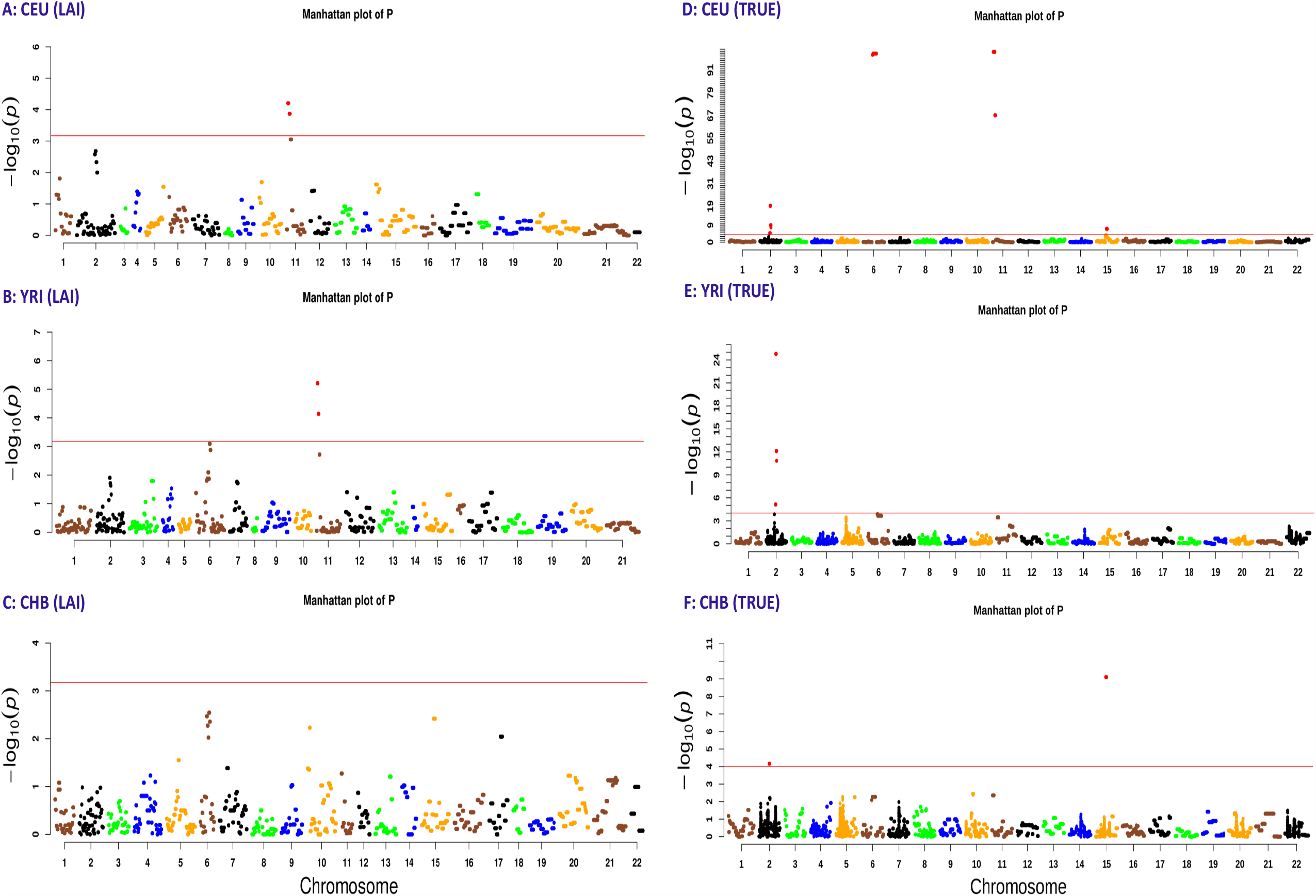
Manhattan plots A, B, and C correspond to the ancestry association analysis using the RFMIX local ancestry inferences (LAI), while D, E, and F correspond to the association using the TRUE local ancestry. A and D correspond to the CEU ancestral population, B and E to the YRI population, and C and F to the CHB population, for the 3-way admixed population analysis. The significance threshold line and the significant SNPs are shown in red.

**Figure 5.**
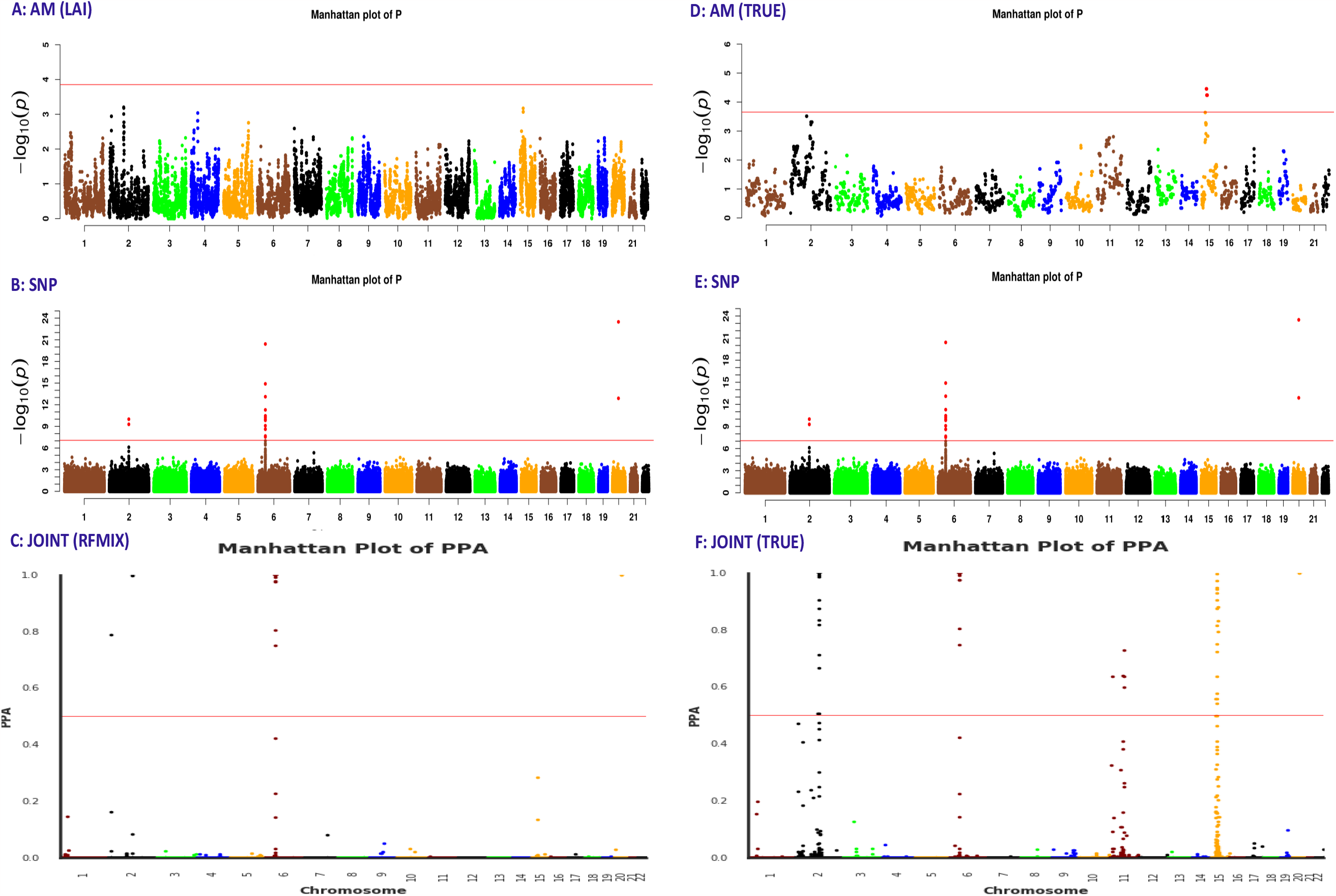
Manhattan plots A, B, and C correspond to the association analysis using the RFMIX local ancestry inferences (LAI), while D, E, and F correspond to the association analysis using the TRUE local ancestry. A and D correspond to the p-values of the MIA per SNP, B and E to the genotype association, and C and F to the joint PPA for the 500 cases and 500 controls in the 5-way admixed population analysis. The significance threshold line and the significant SNPs are shown in red.

**Figure 6.**
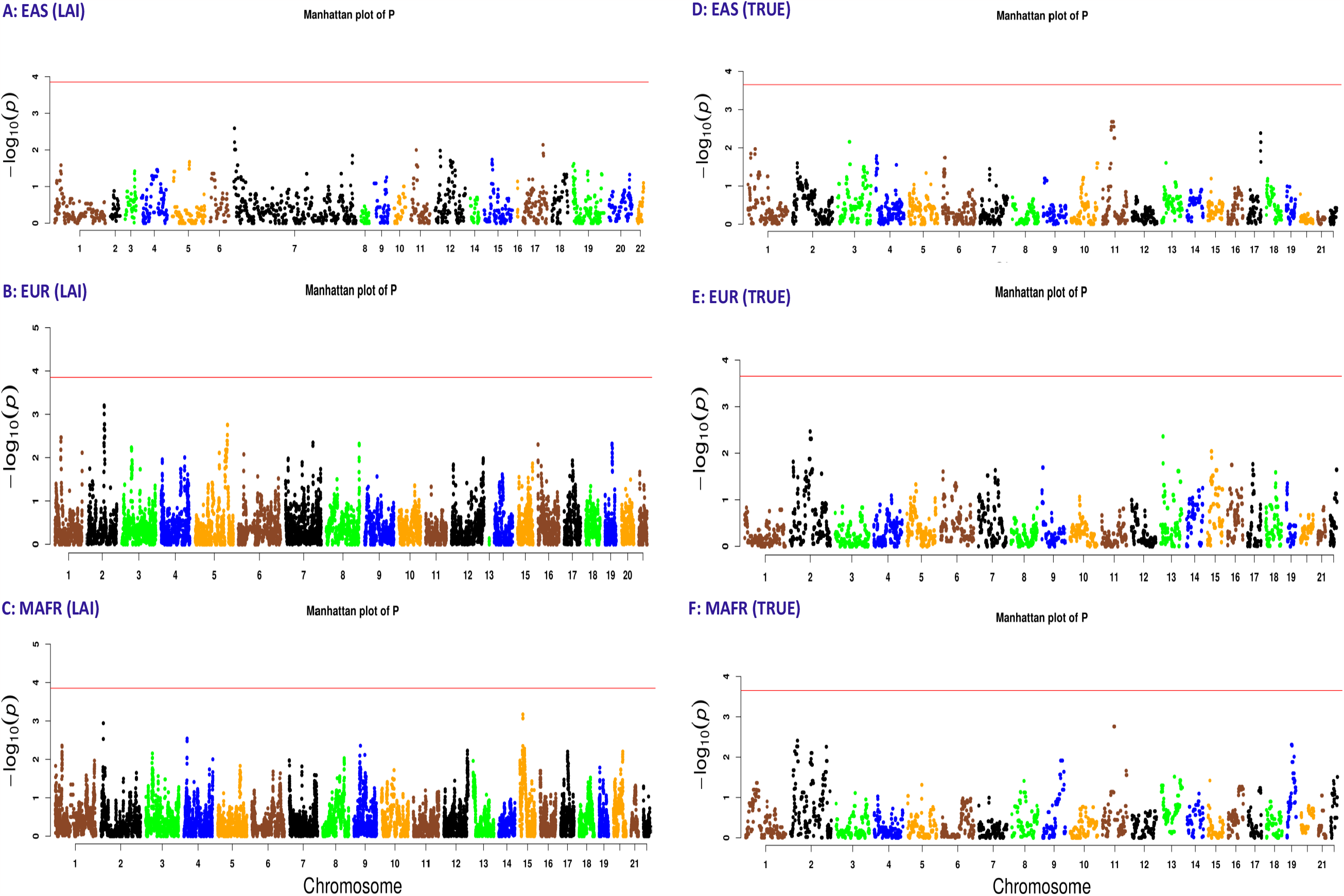
Manhattan plots A, B, and C correspond to the ancestry association analysis using the RFMIX local ancestry inferences (LAI), while D, E, and F correspond to the association using the TRUE local ancestry. A and D correspond to the EAS ancestral population, B and E to the EUR population, and C and F to the MAFR population, for the 500 cases and 500 controls in the 5-way admixed population analysis. The significance threshold line and the significant SNPs are shown in red.

**Figure 7.**
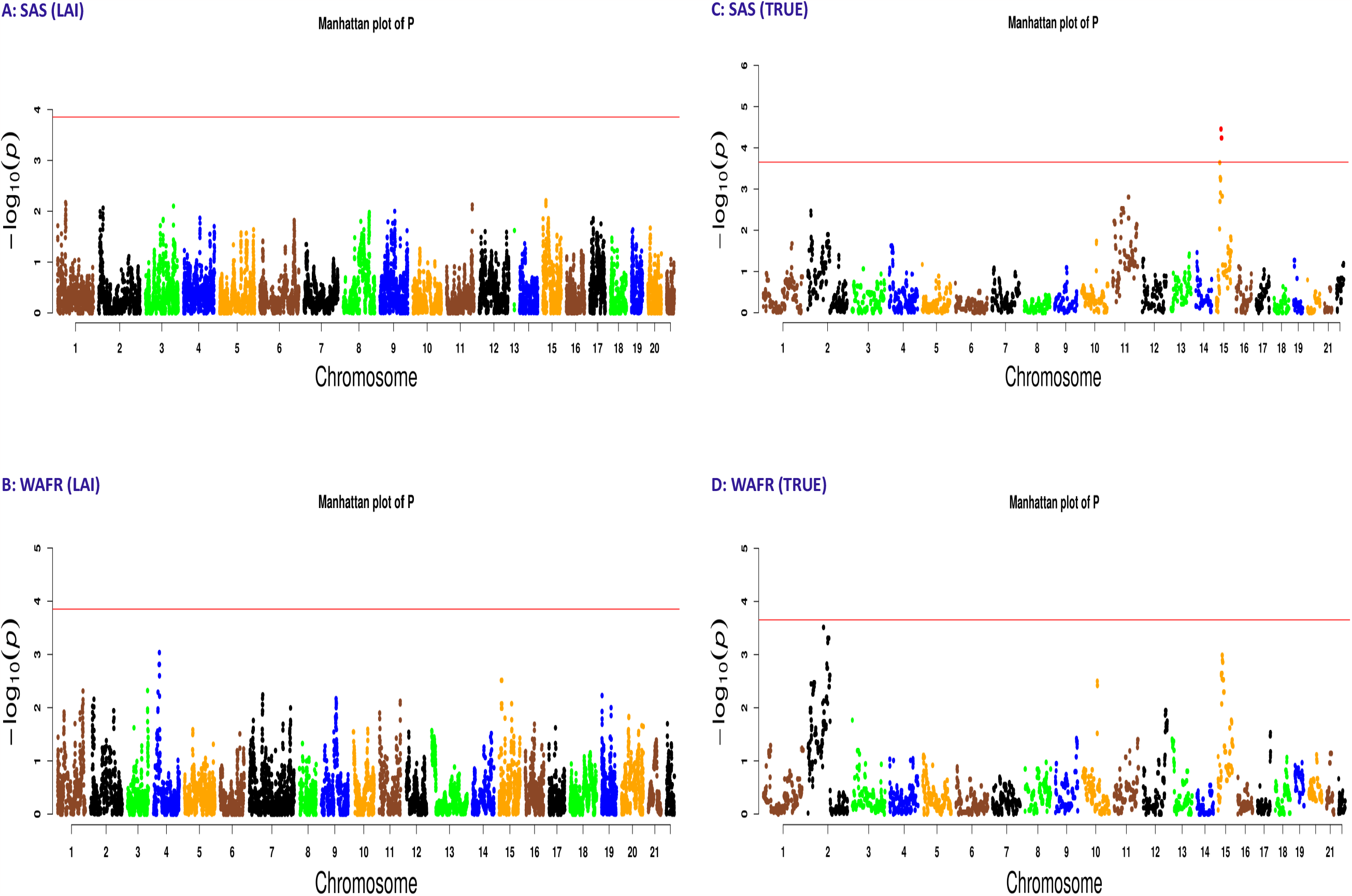
Manhattan plots A and B correspond to the ancestry association analysis using the RFMIX local ancestry inferences (LAI), while C and D correspond to the association using the TRUE local ancestry. A and C correspond to the SAS ancestral population, while B and D correspond to the WAFR population, for the 500 cases and 500 controls in the 5-way admixed population analysis. The significance threshold line and the significant SNPs are shown in red.

**Figure 8.**
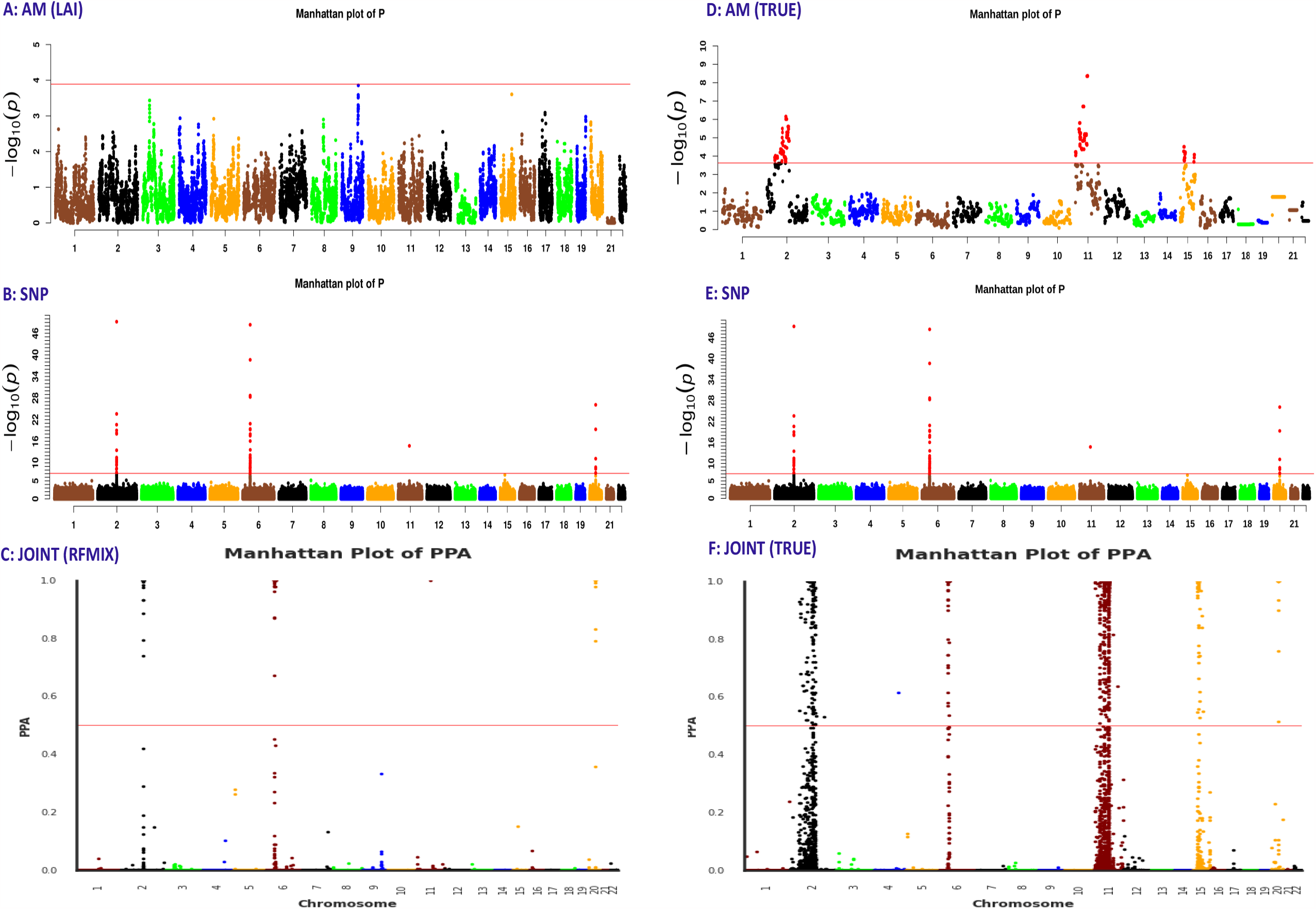
Manhattan plots A, B, and C correspond to the association analysis using the RFMIX local ancestry inferences (LAI), while D, E, and F correspond to the association analysis using the TRUE local ancestry. A and D correspond to the p-values of the MIA per SNP, B and E to the genotype association, and C and F to the joint PPA, for the 2500 cases and 2500 controls in the 5-way admixed population analysis. The significance threshold line and the significant SNPs are shown in red.

**Figure 9.**
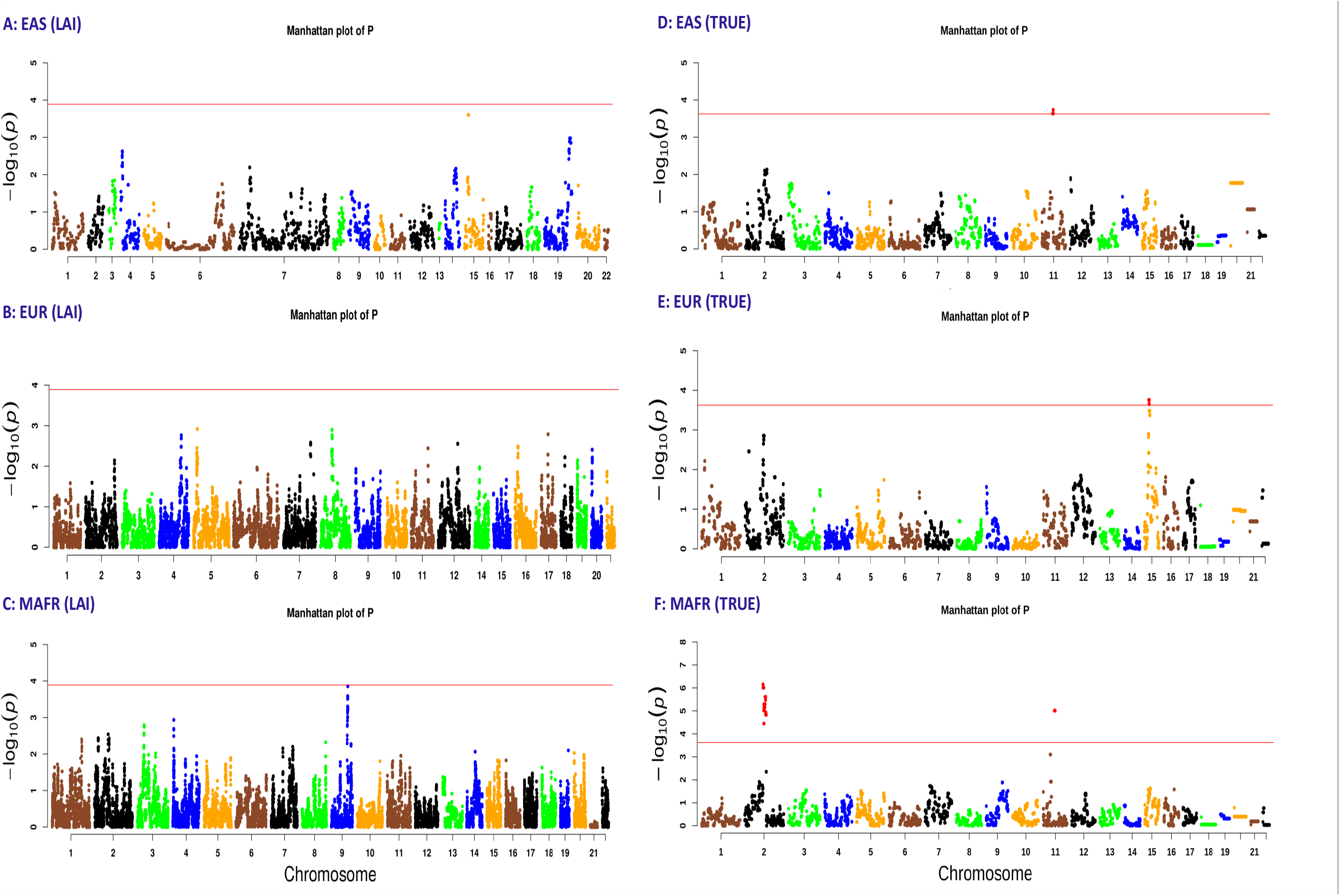
Manhattan plots A, B, and C correspond to the ancestry association analysis using the RFMIX local ancestry inferences (LAI), while D, E, and F correspond to the association using the TRUE local ancestry. A and D correspond to the EAS ancestral population, B and E to the EUR population, and C and F to the MAFR population, for the 2500 cases and 2500 controls in the 5-way admixed population analysis. The significance threshold line and the significant SNPs are shown in red.

We observed from the 3-way simulated admixed population results that by using the RFMIX local ancestry inferences to perform ancestry association, the risk SNP on chromosome 11 was detected as significant; however, all the other simulated signals at chromosomes 2, 6, and 15 were detected at marginal significance thresholds. Of note is that the regions on these chromosomes containing the four risk SNPs were simulated with strong ancestry risk. We also noted that, the genotype association detected the risk SNPs on chromosomes 2 and 6 as significant but those on chromosomes 11 and 15 at marginal or nominal p-values. However, the joint association implemented in JasMAP leveraged the ancestry signals and detected all the simulated risk SNPs at significant thresholds. By using the true local ancestry, we observe that all the simulated risk SNPs were captured at significant levels by the ancestry association analysis, and the joint association was able to capture all the simulated risk SNPs at significant levels. These results indicate that accurate local ancestry boosts the association’s power.

In the 5-way simulated admixed population with the ancestry association and the smaller sample size, we observed that by using the true local ancestry, we were able to observe the simulated risk SNP on chromosome 15 as significant in the ancestry association, while those on chromosomes 2 and 11 were captured at the marginal/nominal level of significance. However, using the RFMIX local ancestry inferences, though the risk SNP on chromosome 15 was captured at a small p-value, it does not reach the significance threshold, and neither did the other simulated risks on chromosomes 2 and 11. The genotype association had significant p-values on chromosomes 2, 6, and 20 and marginal significance in risk SNPs on chromosomes 11 and 15. By applying the joint association to this dataset, we observed that, using the true local ancestry association, we did detect all the simulated risk SNPs as significant except one of the SNPs on chromosome 11, which was detected at a marginal PPA. By using the RFMIX inferences, the joint association was able to capture the risk SNPs on chromosomes 2, 6, and 20 as significant but not those on chromosomes 11 and 15. Of note is that the risk SNPs on chromosomes 2, 6, and 20 have been simulated with a strong genotype signal. In the absence of a strong prior from the ancestry risk on chromosomes 11 and 15 from the ancestry association, the joint association was limited and could only capture the risk SNP on chromosome 15 at a marginal PPA but was not significant.

On the larger 5-way admixed population simulation, we note a similar trend where, using the RFMIX local ancestry inference, none of the simulated risk SNPs in the ancestry association attained the significance threshold, but due to the strong genotype association, the risk SNPs on chromosomes 2, 6, and 20 were detected as significant. Due to an increase in power as a result of increased sample size, one of the SNPs on chromosome 11 was detected as significant in the genotype association and thus also detected as significant in the joint association, irrespective of the weak prior. Using the simulated true local ancestry, we observe that all the risk SNPs on chromosomes 2, 11, and 15 were detected as significant in the ancestry association and thus were also detected as significant in the joint association, irrespective of one risk SNP being detected at marginal significance in the genotype association. The risk SNPs on chromosomes 6 and 20 with strong genotype risk were also detected as having significance in the joint association, irrespective of the absence of ancestry risk using true local ancestry.

We also observed that on chromosome 1 in the 3-way simulation analysis (using the RFMIX local ancestry inferences) and chromosome 4 in the larger 5-way admixed simulation analysis (using the true local ancestry), the joint analysis on JasMAP detected one significant SNP on each of the chromosomes. On further investigation of these two SNPs, we realize that due to random mating simulation in FractalSIM, these two attained marginal significance thresholds in the ancestry association and in the genotype association and were thus detected as significant in the joint association. **Table 8** indicates the summary statistics of these two SNPs. We also observed that with a larger sample size (2500 cases and 2500 controls) in both the 3-way and 5-way analyses, we had quite a substantial increase in the power to detect the risk SNPs in the ancestry association using the true local ancestry, and a lot of SNPs were captured by admixture LD. This was also reflected in the joint association, where the strong ancestry signal empowered a substantial number of SNPs that were detected as significant. We also noted that the ancestry-only association for the 5-way admixed population analysis for the smaller and larger sample size resulted in different MIA for the some of the risk SNPs, but since these were 2 separate simulations, the MIA was determined by MAF of the risk SNP, the relative risks, random mating and mutation simulation.

**Table 8:**
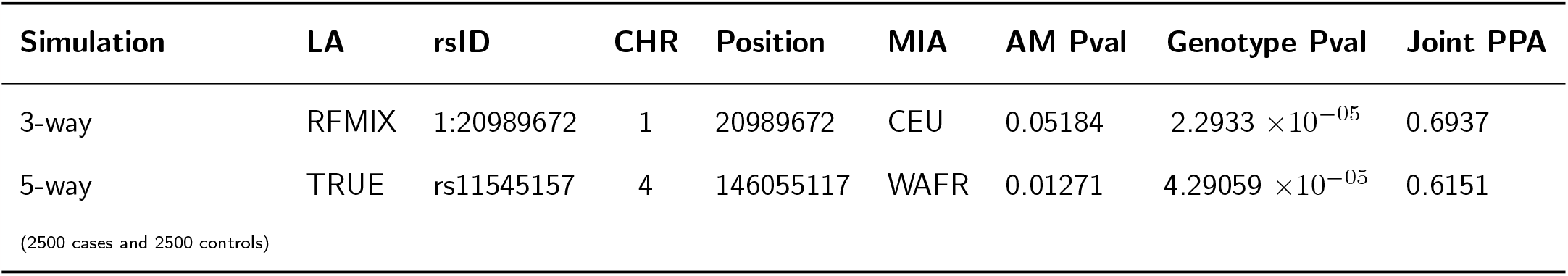
The list of SNPs observed as significant under the joint association (PPA*≥*0.5) on chromosome 1 in the 3-way admixed data analysis and on chromosome 4 in the 5-way admixed (2500 cases, 2500 controls) analysis, with the correponding source of local ancestry (LA), the MIA selected, the p-value under the ancestry-only association (AM Pval), and the genotype-only association (Genotype Pval).

Our general observation in this assessment was that ancestry-only association was only able to detect the risk region where the ancestry risk was strong, irrespective of the presence of a genotype signal. It was also underpowered in regions that had strong genotype signals and no deviations in ancestry. We also observed that the performance of the ancestry association was highly dependent on the local ancestry inferencing, which determined the strength of the prior and consequently the performance of the joint association. The genotype-only association in JasMAP was also robust in detecting regions where the genotype signal was very strong; however, it missed present risk variants when the risk was very weak, and though increasing the sample size increased the power to capture some of the risk variants, others could not reach the stringent genome-wide threshold. The joint approach was able to detect the risk SNPs where the genotypes and ancestry signals were strong. In addition, it was successful in leveraging both the ancestry and genotype signals and improving the power to detect the risk where either approach was underpowered. Of note also was that, though RFMIX had been shown to perform very well in ancestry deconvolution in recently admixed multi-way populations, it struggled to capture the ancestry deviation between the cases and controls at the risk SNP regions but performed better at capturing the deviation in the 3-way admixed population compared to the 5-way admixed population.

### JasMAP Application to a South African Coloured Population

#### Methods

We then applied JasMAP to study the association of tuberculosis (TB) in a South African population commonly referred to as South African Coloured (SAC) located in the Western Cape Province of South Africa, which has a very high incidence of TB. SAC has been shown in previous studies to have five ancestral populations, namely Europeans, Indians, Khoisan, Bantu speakers, and Southeast Asians (Chimusa et al., 2013, 2014).

The study was granted ethics approval by Health Science ethics committees at Stellenbosch University and the University of Cape Town. The cases in the study were identified via bacteriological tests (smear and/or culture positive), while the controls were identified from the same region with no previous history of TB disease or treatment for TB, and the samples were genotyped using an Affymetrix 500 K chip in genome build 36. Association studies of TB in this dataset have been conducted previously using other GWAS tools (Chimusa et al., 2014).

We performed quality control on the data by selecting only SNPs with MAF*>* 0.01, excluding individuals with missing genotypes *>* 0.05%, performing the Hardy-Weinberg equilibrium test in controls at *α* = 0.0001 and implementing PLINK to remove related individuals. This resulted in a dataset of 733 samples, with 642 cases and 91 controls and a total of 272,796 autosomal SNPs. We further removed 37,556 A/T and C/G and 6,176 sex SNPs, which further reduced our dataset to 272,076 SNPs.

We lifted over our dataset from genome build 36 to genome build 37, using the liftOver tool provided freely online by the Centre for Statistical Genetics at the University of Michigan. We then performed imputation on our dataset using the Sanger Imputation Service (Das et al., 2016) by the Wellcome Sanger Institute, also freely available online, where we chose the African Genome Resource as our reference. We then performed post-imputation quality control on the dataset and removed all the SNPs with MAF*<* 0.05 and genotypes missingness *>* 0.02. We also checked for individual missingness, and all the samples had *<* 0.02 missingness. Our final number of SNPs was 6,055,402.

We used proxy ancestral populations to deconvolve SAC ancestry, including CEU, CHB, GIH, KHS, and YRI, which were publicly available datasets. CEU, CHB, and YRI were described in **Table 1**, while the GIH were 109 Gujarati Indians from Houston, Texas, USA, obtained from 1000 genomes, and the KHS were 24 Khoisan from South Africa, publicly available in the Human Genome Diversity Project (HGDP) and Schlebusch et al. (2012). We then obtained shared SNPs in all six populations, where we had a total of 5,617,504 SNPs, and implemented RFMIX to perform local ancestry inferencing. We first estimated the global ancestry using the ADMIXTURE tool and also by averaging the RFMIX local ancestry inferences per individual.

We then implemented GCTA and obtained the first 10 PCs, which we used as our covariates to account for the global ancestry. We ran our custom script to obtain the effective number of tests for SAC, which we obtained as 1294 tests with a corresponding ancestry significance threshold of 3.86399 *×* 10^*−*05^. We then ran JasMAP similar to the simulated datasets and also performed association analysis using GCTA and SNPTEST-Bayesian for comparison purposes and obtained the corresponding Manhattan plots.

Further, we extracted all SNPs that had a PPA of ⩾ 0.05 and performed functional annotation of the SNPs and gene mapping analysis using the FUMA (Watanabe et al., 2017) tool, which is also freely available online. Since FUMA accepts p-values as input, we used our genotype summary statistics as input for the SNP and listed the SNPs that had PPA ⩾ 0.5 as the lead SNPs.

## Results

**Figure 11** shows the admixture plots of the global ancestry estimates from ADMIXTURE and RFMIX. The global ancestry estimates from RFMIX were CEU - 7%, CHB - 3%, GIH - 5%, KHS - 58%, and YRI - 27%, while those from the ADMIXTURE tool were CEU - 2%, CHB - 1%, GIH - 6%, KHS - 89%, and YRI - 2%. We observed that the estimates from RFMIX were closer to the estimates from previous studies of the SAC population that had estimated the global ancestries as 19*±*1% from CEU, 7*±*0.5% from CHB+JPT, 13*±*0.9% from GIH, 33*±*1% from KHS, and 28*±*2% from YRI (Chimusa et al., 2014).

**Figure 10.**
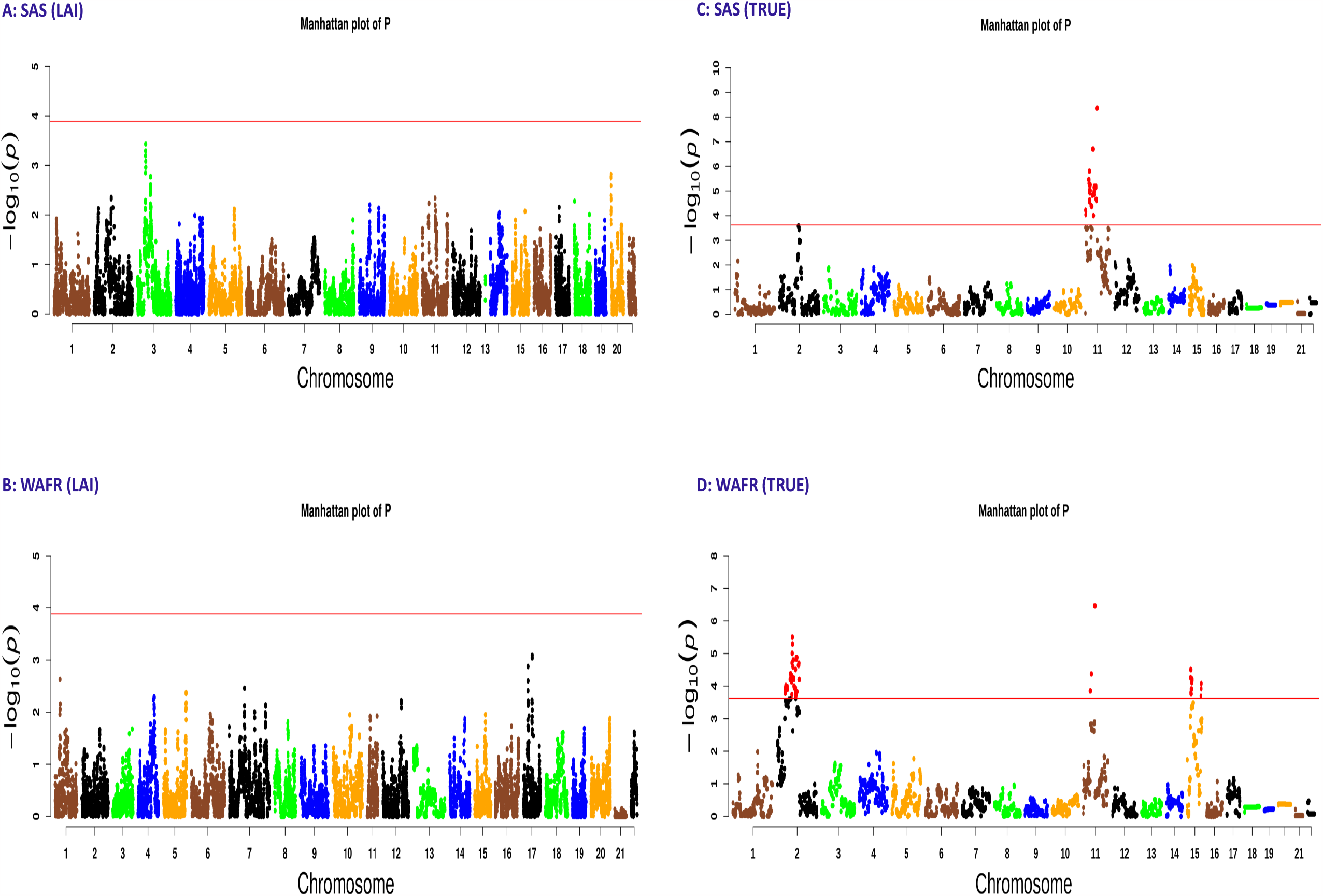
Manhattan plots A and B correspond to the ancestry association analysis using the RFMIX local ancestry inferences (LAI), while C and D correspond to the association using the TRUE local ancestry. A and C correspond to the SAS ancestral population, while B and D correspond to the WAFR population, for the 2500 cases and 2500 controls in the 5-way admixed population analysis. The significance threshold line and the significant SNPs are shown in red.

**Figure 11.**
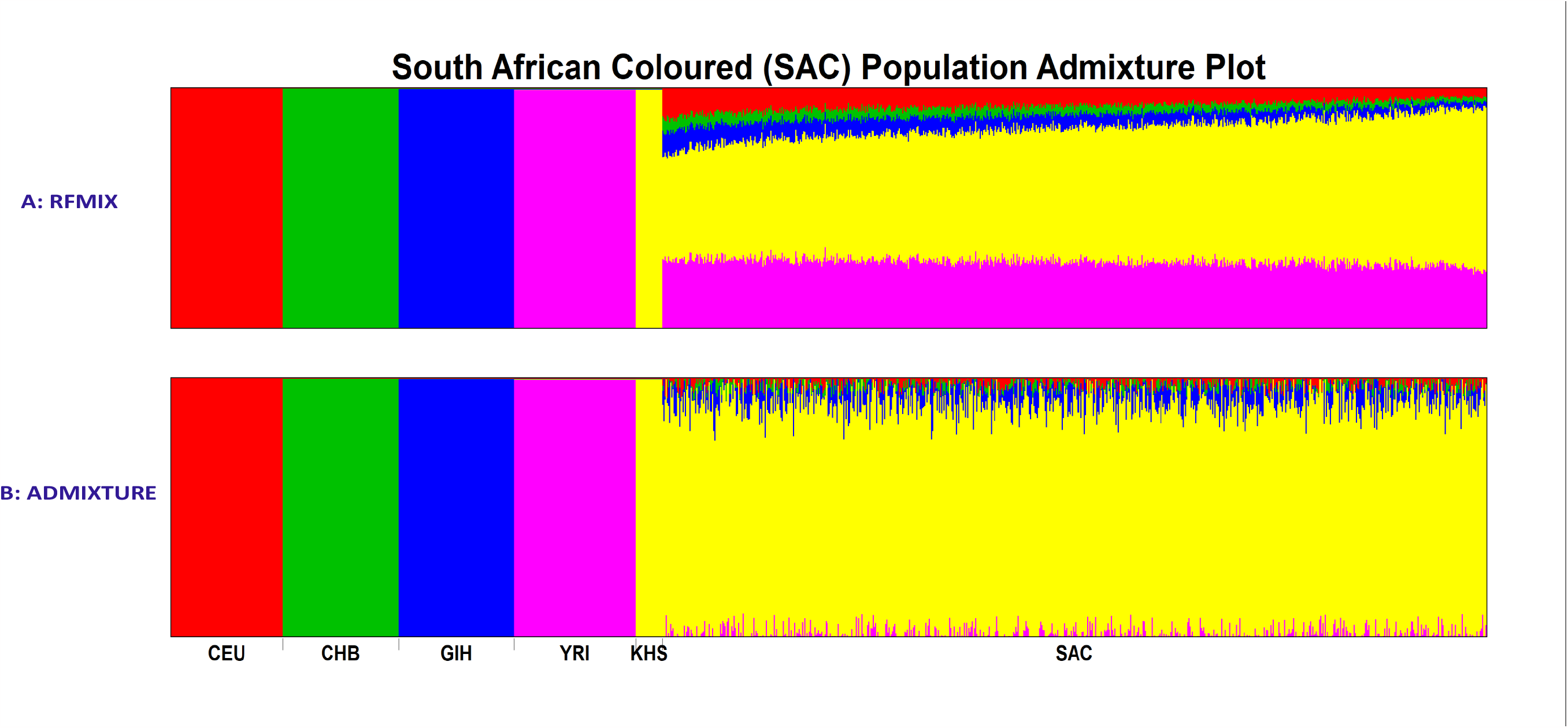
The admixture plot for of the SAC population, and the corresponding CEU, CHB, GIH, YRI, and KHS reference populations used as proxy ancestral populations. A: is the admixture block estimated by the local ancestry averages from RFMIX and B: is the admixture block of the global ancestry proportion estimated by ADMIXTURE tool.

Our ancestry association identified a region on chromosome 9 that was significant; however, the JasMAP genotype association, similar to both the GCTA and SNPTEST association tests, did not detect any SNP as significant. The joint association test on JasMAP detected 13 SNPs with PPA⩾ 0.5 that were detected either at marginal or significant thresholds in the ancestry and at marginal thresholds in the genotype association. We extracted 100 SNPs that had a PPA of ⩾ 0.05 for the functional annotation and gene mapping, and the 13 SNPs listed in **Table 9** were listed as the lead SNPs. 9 of the SNPs we detected as significant were identified as intergenic, 2 as intronic, and the remaining 2 as non-coding RNA intronic. Gene mapping analysis reported genes that were close to our significant SNPs, and these have been listed in **Table 9**.

**Table 9:** A list of SNPs detected as significant in the joint association (PPA *≥* 0.5) with the corresponding p-values for the ancestry-only association (AM Pval) and genotype-only association (Genotype Pval) for the association analysis of the SAC population, with corresponding function and nearest gene.

| rsID | CHR | Position | Ancestry | AM Pval | Genotype Pval | Joint PPA | Function | Nearest Gene |
| --- | --- | --- | --- | --- | --- | --- | --- | --- |
| rs113164717 | 2 | 8836489 | GIH | 0.00442 | $1.28506 \times 10^{-05}$ | 0.7163 | intergenic | ID2 |
| rs17050321 | 4 | 139285676 | CEU | 0.01712 | $1.54109 \times 10^{-06}$ | 0.8057 | ncRNA_intronic | LINC00499 |
| rs9375980 | 6 | 133995787 | GIH | $4.659 \times 10^{-05}$ | 0.00054 | 0.7907 | ncRNA_intronic | RP3-323P13.2 |
| rs16917540 | 9 | 115698356 | CHB | 0.00053 | $7.32944 \times 10^{-05}$ | 0.8166 | intergenic | MUPP |
| rs10817412 | 9 | 115702989 | CHB | 0.00053 | $8.4919 \times 10^{-05}$ | 0.7810 | intergenic | MUPP |
| rs9517023 | 13 | 98374272 | CEU | 0.00065 | 0.00016 | 0.5091 | intergenic | RP11-120E13.1 |
| rs9517027 | 13 | 98376647 | CEU | 0.00065 | 0.00016 | 0.5091 | intergenic | RP11-120E13.1 |
| rs9517028 | 13 | 98377240 | CEU | 0.00065 | 0.00016 | 0.5091 | intergenic | RP11-120E13.1 |
| rs9517032 | 13 | 98383019 | CEU | 0.00065 | 0.00016 | 0.5091 | intergenic | RP11-120E13.1 |
| rs9517034 | 13 | 98383079 | CEU | 0.00065 | 0.00016 | 0.5091 | intergenic | RP11-120E13.1 |
| rs4570837 | 16 | 60140710 | YRI | 0.00255 | $2.40286 \times 10^{-05}$ | 0.7113 | intergenic | RP11-430C1.1 |
| rs9931450 | 16 | 76566769 | KHS | 0.00165 | $2.48788 \times 10^{-06}$ | 0.9900 | intronic | CNTNAP4 |
| rs9921377 | 16 | 76594727 | KHS | 0.00165 | $4.56263 \times 10^{-05}$ | 0.6496 | intronic | RP11-58C22.1 |

By gene pathway search on the Kyoto Encyclopedia of Genes and Genomes (KEGG) database (Kanehisa et al., 2023), gene *ID2*, inhibitor of DNA binding 2, was associated with the TGF-beta and the Hippo signaling pathways. Gern et al. (2021), in their publication, found that the TGF-beta signaling pathway was linked to cell suppression for local CD4 T cells in the tuberculous granuloma, while Boro et al. (2016) found that the Hippo signaling pathway was linked to the modulation of the host immune response to bacterial infections. Smoking tobacco has been noted in several studies (Davies et al., 2006; Alavi-Naini et al., 2012; Chakaya et al., 2021) to lead to an increased risk of TB. Saunders et al. (2022), in a recent study, investigated genetic associations to alcohol and tobacco consumption in diverse populations and linked the *ID2* gene to regular tobacco smoking. *LINC00499*, long intergenic non-protein coding RNA 499, and *RP11-430C1*.*1*, also known as long intergenic non-protein coding RNA 2141 (*LINC02141*), have been reported by both Liu et al. (2019) and Saunders et al. (2022) on the GWAS catalog to also be linked to smoking initiation. *CNTNAP4* is also another gene that has been linked to smoking behavior and reported on the GWAS catalog by Meddens et al. (2019). The Ensembl database (Martin et al., 2023) reports gene *RP11-58C22*.*1* as a putative protein coding that has been retired, and *CNTNAP4* was the next closest gene to SNP *rs*9921377 as reported by FUMA. *MUPP*, the major urinary protein pseudogene, has no ability to produce a functional protein and is the only major urinary protein in humans (Charkoftaki et al., 2019). The next closest gene to SNPs *rs*10817412 and *rs*16917540 on chromosome 9 was *RP11-4O1*.*2*, also known as long intergenic non-protein coding RNA 2977 (*LINC02977*), which has been reported by Comuzzie et al. (2012) on the GWAS catalog to be associated with obesity. Badawi et al. (2020), in a recent analysis on the relationship between TB and obesity, identified obesity as a protective factor for TB. Although gene *RP3-323P13*.*2*, also known as Tcf21 antisense RNA inducing demethylation (*TARID*), has been linked to lung cancer by several studies (Arab et al., 2014; Shi et al., 2017) and the association is also reported in the GWAS catalog by Ji et al. (2018), the gene has no reported link to TB in the literature. However, its interaction with the growth arrest and DNA damage-inducible alpha (*GADD45A*) gene has been identified in several studies (Arab et al., 2014, 2019). Daya et al. (2014) in an ancestry-only association study of a South African population with TB found *GADD45A* to be close to a region that had an excess of San ancestry in their cases. The KEGG database associates the *GADD45A* gene with the NF-kappa B signaling pathway, which has been shown to both aid mycobacterium tuberculosis infection and also support immune function to clear the infection (Poladian et al., 2023). Possible association of *TARID* to TB via epistastic is an area of investigation in future studies. *RP11-120E13*.*1* is a long intergenic noncoding RNA whose function or pathways are yet to be reported in the literature and is thus another candidate for future investigation on a possible link to TB.

We also checked all genes that were 100 kb away from the significant SNP, and we found *SLC7A11*, a protein-coding gene (also known as *xCT*), was close to SNP *rs*17050321 on chromosome 4. Wang et al. (2020) found *SLC7A11* to be linked to an increased susceptibility to TB in the Chinese population. SNP *rs*7624965 on chromosome 3, detected with a PPA of 0.164, was included in the functional annotation and was close to a protein-coding gene, *MASP1*, which Klassert et al. (2018) found to be associated with the development of TB in a study of an Indian population. To our knowledge, none of the other detected genes have been linked to TB.

## Discussion

Extension of GWAS to diverse populations is imperative, but unfortunately, association studies in these populations are still haunted by small sample sizes. An approach that leverages other features of these populations to improve association power is thus vital. In this study, we have developed a joint association analysis tool, JasMAP, that leverages ancestry and genotype risk to improve the power to detect risk variants in association studies. Through simulation of case-control 3-way and 5-way admixed indiviuals, we were able to demonstrate that JasMAP is robust and performs better than current commonly used tools in detecting present-risk SNPs in association analysis of multi-way admixed populations, particularly where the genotype risk is very weak and there is an ancestry risk. We have demonstrated that JasMAP is able to increase power in studies with small sample sizes.

We also applied JasMAP joint association to a TB phenotype, where we identified 13 significant SNPs. 12 of these were not identified by the genotype-only or ancestry-only associations, and 1 SNP was identified as significant in the ancestry association but not in the genotype association. Through functional annotation of the significant SNPs, we found the SNPs to be either intergenic or located in an intron. By gene mapping, these SNPs were linked to 8 novel genes by positional mapping, 5 of which are functionally linked to TB, while the remaining 3 could further be investigated for possible links to the disease. We also replicated *SLC7A11* and *MASP1* genes that have previously been linked to TB.

In addition to the joint association, JasMAP outputs the ancestry-only and genotype-only association summary statistics to the user. An additional script to calculate the effective number of tests for the local ancestry inferences is provided, and the other input formats for the required datasets in JasMAP are easy to obtain. JasMAP is implemented in the Python language, very user-friendly, and publicly available for use (https://github.com/JQ-Mugo/JasMAP).

Our joint approach, however, does have some limitations. The JasMAP joint approach is highly dependent on the accuracy of local ancestry inferencing. We observed in our analysis that one of the best available tools to deconvolve ancestry in multi-way admixed populations in recently admixed populations, RFMIX, performed fairly well in detecting ancestry deviation between the cases and controls in 3-way admixed populations when the ancestry risk was very strong. However, RFMIX was limited in detecting in the deviation in the 5-way admixed population, which affected the joint analysis. However, local ancestry deconvolution in multi-way admixed populations is still an ongoing problem of interest to many researchers, and more robust tools are being developed.

Our LMM approach currently assumes that the genetic matrix *K* is full rank. Extension of our method to lower-rank approaches is theoretically possible, but we have left that for a future update of JasMAP. We have assessed JasMAP using a binary phenotype, but JasMAP can be applied to a quantitative phenotype.

We have tested JasMAP on 3-way and 5-way admixed populations, but the tool can be run for any *K >* 1 number of ancestries. Though the 2-step approach in JasMAP implies it is relatively slower than the other association tools, it can be run on a computer cluster with the option to parallize the analysis to mitigate the computational cost.

## Statements and Declarations

## Data Availability

The reference data used in this study is publicly available in the 1000 Genomes resources, Human Genome Diversity Project (HGDP), and from Schlebusch et al. (2012). Simulated data is also available by request from the corresponding author.

## Acknowledgments

We acknowledge Prof. Marlo Moller, who provided part of the data used in this research work, and the study participants involved in this study. We acknowledge the High Performance Computing resources at the University of Cape Town and the National Integrated CyberInfrastructure System (NICIS)-Centre for High Performance Computing for providing access to the computer clusters that were used to run all the analysis in this study.

## Data Availability

The reference data used in this study is publicly available in the 1000 Genomes resources, Human Genome Diversity Project (HGDP), and Schlebusch et al. (2012). Simulated data is also available by request from the corresponding author.

## Conflict of interest

There are no conflicts to declare.

The Manhattan plots for the MIA per SNP, the genotype association, and the joint PPA are shown in **Figure 12**, while the ancestry-specific Manhattan plots of the ancestry association are shown in **Figure 13**. **Figure 14** shows the Manhattan plots for the GCTA and SNPTEST association analysis, while the summary statistics of the SNPs that obtained a PPA⩾ 0.5 are listed in **Table 9**.

**Figure 12.**
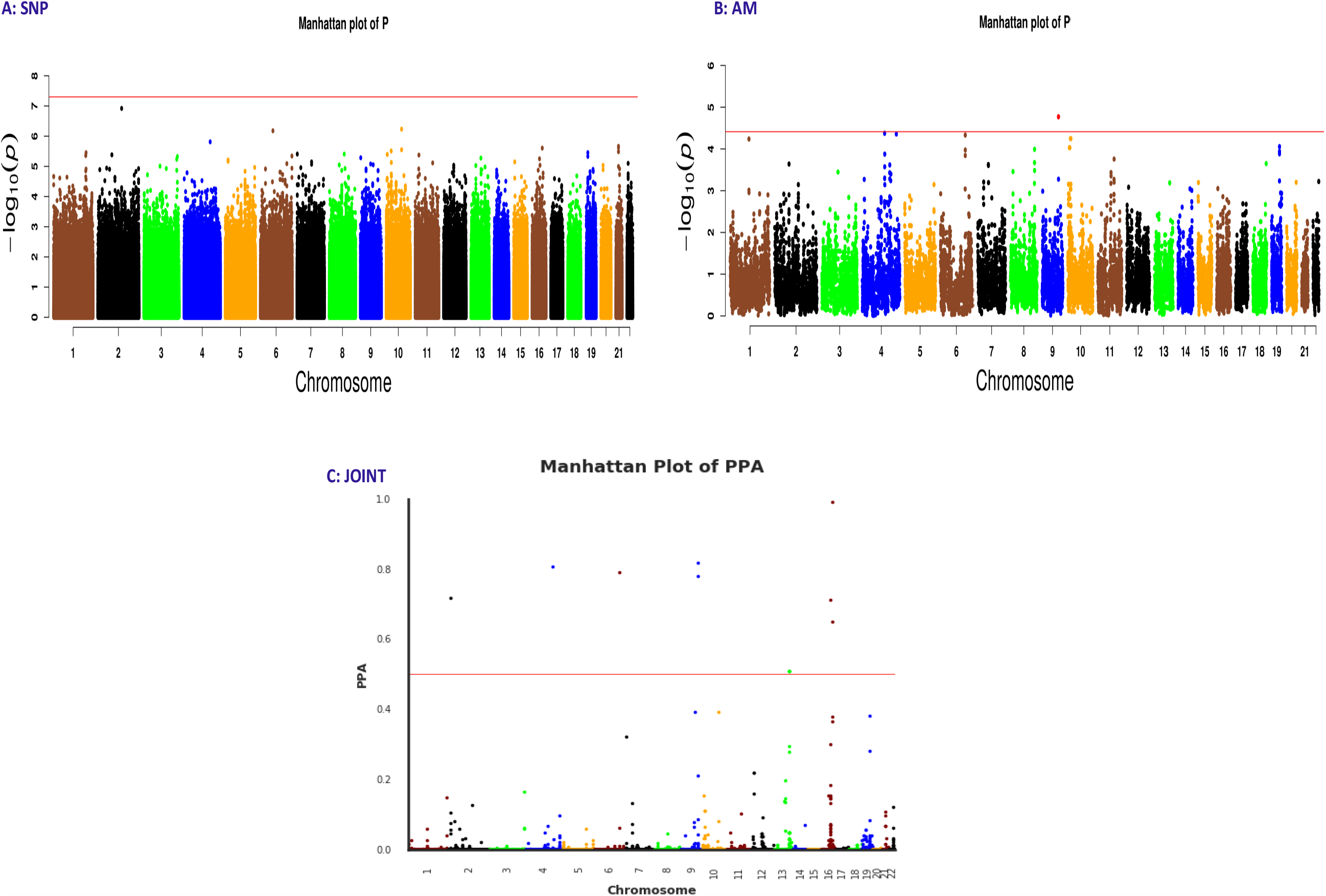
The Manhattan plots A and B correspond to genotype-only association and the p-values of the MIA per SNP, respectively for the association tests of SAC population, while C is the joint PPA Manhattan plot. The significance threshold line and the significant SNPs are shown in red.

**Figure 13.**
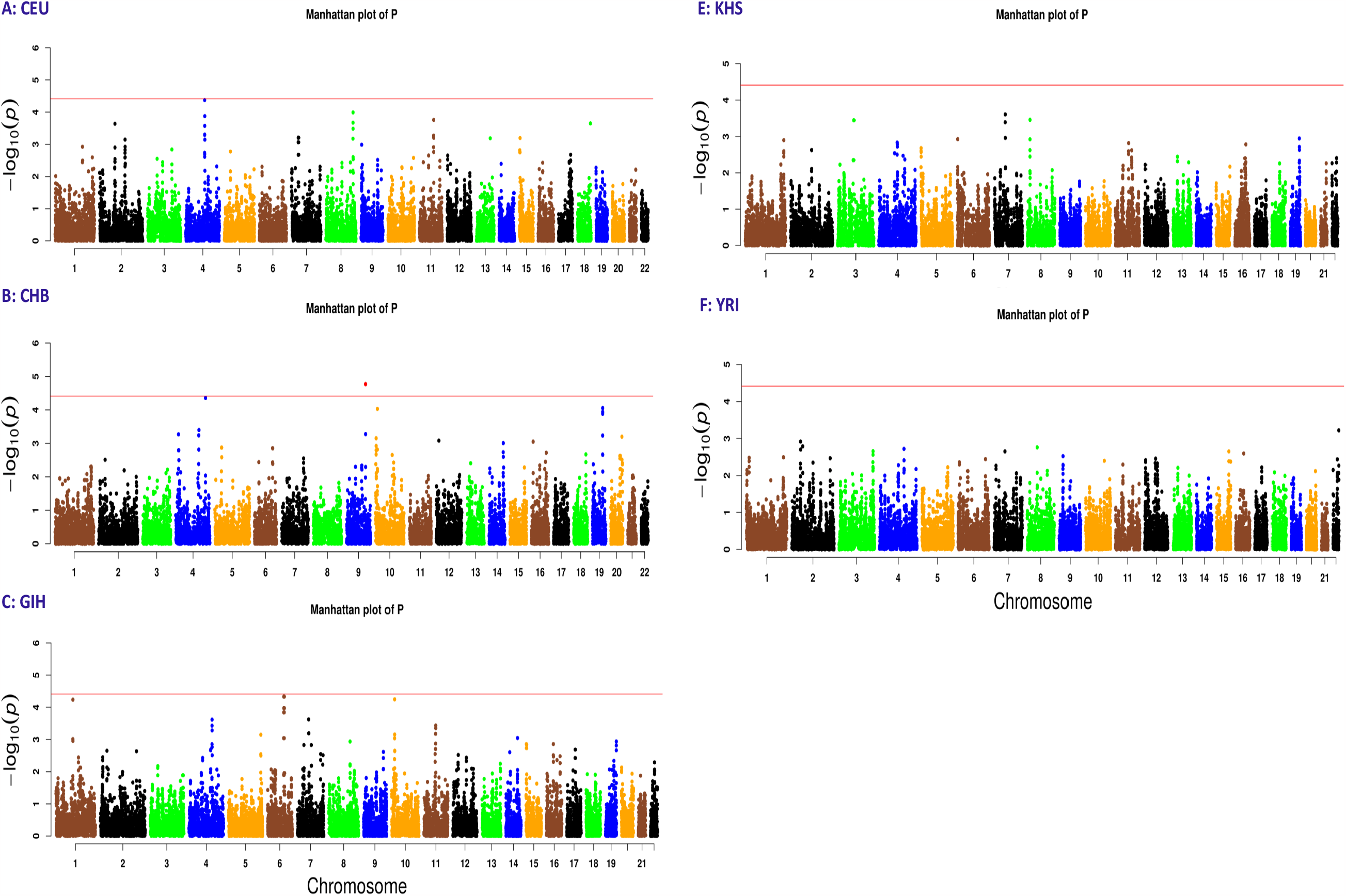
The Manhattan plots of the ancestry-only association analysis using the RFMIX local ancestry inferences (LAI), for A: CEU, B: CHB, C: GIH, D: KHS and E: YRI ancestral populations. The significance threshold line and the significant SNPs are shown in red.

**Figure 14.**
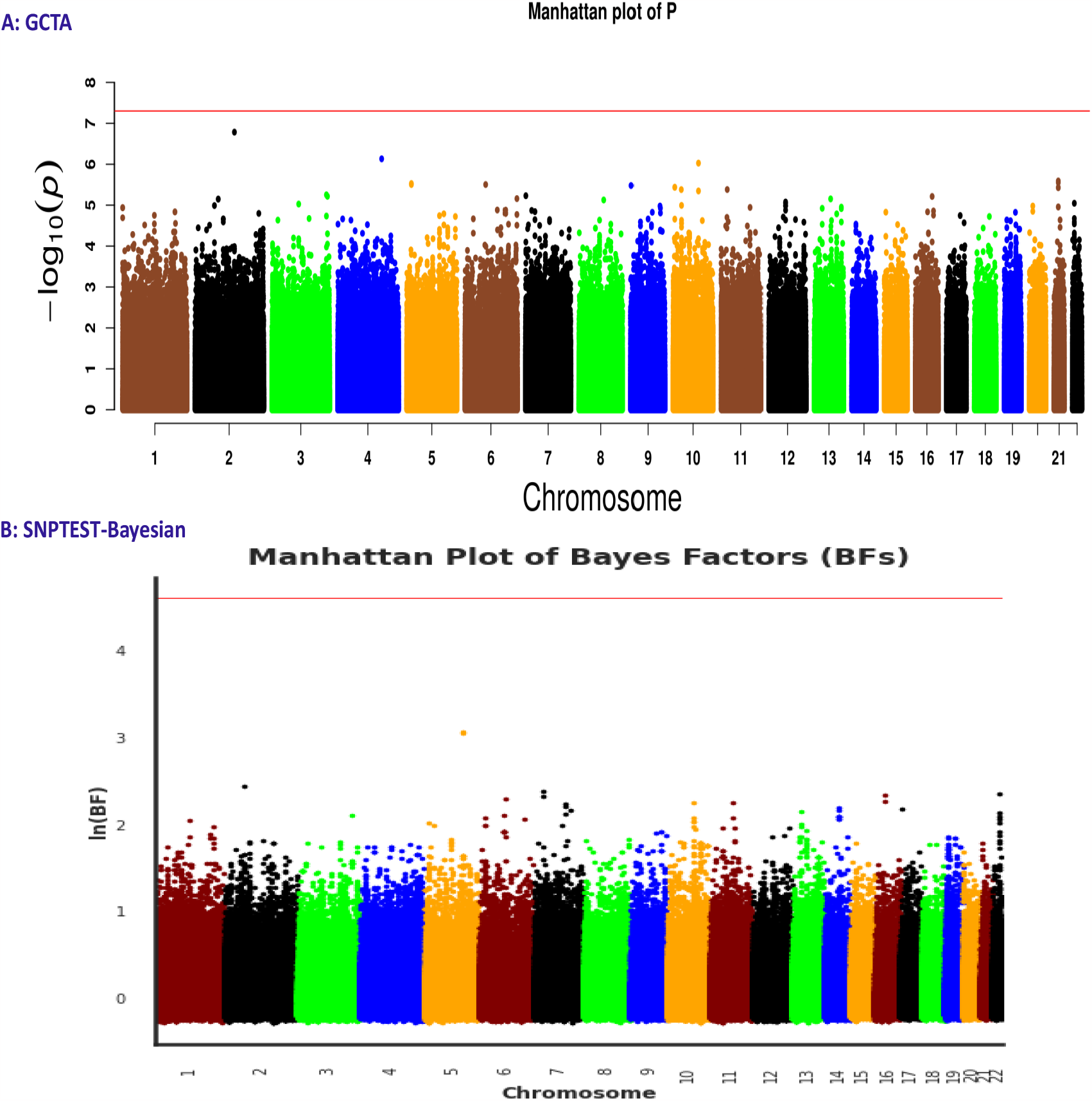
The Manhattan plots of the genotype-only association analysis using A: GCTA and B: SNPTEST-Bayesian GWAS tools. The significance threshold line and the significant SNPs are shown in red.

